# Changing Agendas on Sleep, Treatment and Learning in Epilepsy (CASTLE) Sleep-E: A protocol for a randomised controlled trial comparing an online behavioural sleep intervention with standard care in children with Rolandic epilepsy

**DOI:** 10.1101/2022.05.31.22275803

**Authors:** CASTLE Group, CASTLE Advisory Panel, Nadia Al-Najjar, Lucy Bray, Bernie Carter, Amber Collingwood, Georgia Cook, Holly Crudgington, Kristina C. Dietz, Paul Gringras, Will A. S. Hardy, Harriet Hiscock, Dyfrig Hughes, Christopher Morris, Hannah Morris, Deb K. Pal, Morwenna Rogers, Alison Rouncefield-Swales, Holly Saron, Catherine Spowart, Lucy Stibbs-Eaton, Catrin Tudur Smith, Victoria Watson, Luci Wiggs, Paula R Williamson, Eifiona Wood

**Affiliations:** King’s College London, UK; CASTLE Research Programme, UK; University of Liverpool, UK; Edge Hill University, UK; Oxford Brookes University, UK; Evelina London Children’s Hospital, UK; Bangor University, UK; Murdoch Children’s Research Institute, Australia; University of Exeter, UK; University of Central Lancashire, UK

**Keywords:** Epilepsy, Rolandic, Sleep, Child, Randomized Controlled Trials as Topic, Evidence-Based Medicine

## Abstract

**Introduction:** Sleep and epilepsy have an established bi-directional relationship yet only one randomised controlled clinical trial has assessed the effectiveness of behavioural sleep interventions for children with epilepsy. The intervention was successful, but delivered via face-to-face educational sessions with parents, which are costly and non-scalable to population level. The Changing Agendas on Sleep, Treatment and Learning in Epilepsy (CASTLE) Sleep-E trial addresses this problem by comparing clinical- and cost-effectiveness in children with Rolandic epilepsy between standard care and standard care augmented with a novel, tailored parent-led CASTLE Online Sleep Intervention (COSI) that incorporates evidence-based behavioural components.

**Methods and analyses:** CASTLE Sleep-E is a UK-based, multi-centre, open label, randomised, parallel-group, pragmatic superiority trial. A total of 110 children with Rolandic epilepsy will be recruited in out-patient clinics and allocated 1:1 to standard care (SC) or standard care augmented with COSI (SC + COSI). Primary clinical outcome is parent-reported sleep problem score (Children’s Sleep Habits Questionnaire). Primary health economic outcome is the Incremental Cost Effectiveness Ratio (National Health Service and Personal Social Services perspective, Child Health Utility 9D instrument). Parents and children (≥ 7 years) can opt into qualitative interviews and activities to share their experiences and perceptions. Jointly, the qualitative trial component and the COSI system (e-analytics and evaluation module) will provide information for a process evaluation (context, implementation, and mechanisms of impact).

**Ethics and dissemination:** The CASTLE Sleep-E protocol was approved by the Health Research Authority East Midlands – Nottingham 1 Research Ethics Committee, reference: 21/EM/0205. Trial results will be disseminated to scientific audiences, families, professional groups, managers, commissioners, and policy makers. Pseudo-anonymised Individual Patient Data will be made available after dissemination on reasonable request.

**Registration details:** ISRCTN registry (Trial ID: ISRCTN13202325, prospective registration 09/Sep/2021). See Supplemental Table 1 for the World Health Organisation Trial Registration Data Set (Version 1.3.1).

**Strengths and limitations of this study:**

- First randomised controlled trial to evaluate the clinical- and cost-effectiveness of a novel, tailored, parent-led CASTLE Online Sleep Intervention (COSI) that incorporates evidence-based behavioural components for children with Rolandic epilepsy
- Extensive Patient and Public Involvement via dedicated CASTLE Advisory Panel
- Embedded health economic evaluation
- Embedded process evaluation
- Limitation: Heavily reliant on participant self-report to assess intervention implementation, ameliorated by COSI e-analytics and actigraphy data

## INTRODUCTION

Epilepsy is one of the most common long-term neurological conditions worldwide; its prevalence peaks during childhood (5–9 years) and again late in life (over 80 years).^1^ Epilepsy in children (5 to <13 years) accounts for the annual loss of over 2.6 million disability-adjusted life years, equivalent to 1.8 % of the global burden of disease from all causes among children and adolescents.^2^ Rolandic Epilepsy (RE) is the most common childhood epilepsy.^3^

In the UK, RE has a stable crude incidence rate of 5 in 100 000 children (<16 years) or 542 new cases annually.^4^ Concurrent neuro-developmental disorders are very common (35 %).^5^ Seizures are often triggered by sleep fragmentation.^6^ Many parents co-sleep or monitor children with nocturnal seizures, and children themselves experience a fear of death during and after a seizure.^7^ Problems related to sleep emerge as one of the top concerns for both children and parents,^8^ but are often unaddressed.^9 10^

A recent systematic review and meta-analysis of clinical trials shows that parent-based behavioural sleep interventions are effective for typically-developing children and those affected by neurological and neuro-developmental disorders.^10^ The review concluded that randomised controlled clinical trials assessing functional outcomes (e.g. cognition, emotion, behaviour) and targeting specific populations (e.g. epilepsy) are missing (but see two recent trials).^11 12^ Harms capture for cognitive-behavioural and behavioural sleep interventions has been sparse (only 32.3 % of trials address Adverse Events) and predominantly inadequate (92.9 % of trials do not meet adequate reporting criteria).^13^ Observed harms of behavioural sleep interventions in adults have been mild (e.g. transient fatigue/exhaustion from sleep restriction in insomnia in 25–33 % of participants).^14^ The only published paediatric and adult epilepsy trials did not address harms.^11 12^ Based on the existing evidence, the benefits of behavioural sleep interventions in children with epilepsy outweigh potential harms, especially because sleep problems not only affect seizure control, but overall child well-being, learning and memory, and parental quality of life.^9 10^ There remains, however, uncertainty whether sleep interventions, which can be resource intensive, are cost-effective in public health systems.

This protocol describes the design for the Changing Agendas on Sleep, Treatment and Learning in Epilepsy (CASTLE) Sleep-E trial, which evaluates the clinical- and cost-effectiveness of a novel, tailored, parent-led CASTLE Online Sleep Intervention (COSI) that incorporates evidence-based behavioural components for children with epilepsy. COSI and CASTLE Sleep-E outcome-selection were co-produced by affected children, young people, and their parents, sleep- and epilepsy experts.^8 15–17^ The CASTLE Sleep-E protocol follows Standard Protocol Items: Recommendations for Interventional Trials (SPIRIT),^18 19^ its extension for Patient Reported Outcomes (SPIRIT-PRO),^20^ and the Guidance for Reporting Involvement of Patients and the Public (GRIPP2).^21^

As CASTLE Sleep-E is a pragmatic superiority trial assessing whether UK standard care for children with RE should be augmented with an online behavioural sleep intervention, standard care is the appropriate comparator.^22–24^ Current UK clinical guidelines^25–27^ recommend that standard care for children with RE consist of a comprehensive care plan with the option of pharmacological treatment with anti-epileptic drugs.

The primary objective of CASTLE Sleep-E is to determine if standard care augmented with COSI is superior to standard care alone in reducing sleep problems in children with RE and cost-effective. Implementation details and secondary objectives are reported in Table 1.

**Table 1.** Outcomes for CASTLE Sleep-E (incl. participant level metrics, time-points, aggregation method). Child measures may be collected by parent proxy.

| Outcome type | Specific measurement variable | Collected for | Participant-level analysis metric | Measurement time-point(s) |
| --- | --- | --- | --- | --- |
| Primary |  |  |  |  |
| Clinical | Children's Sleep Habits Questionnaire | Child | Total score | Baseline<br>3 months |
| Health economic | Health utility<sup>1</sup> | Child<br>Parent | Time integral of utility | Baseline<br>3 months<br>6 months |
| Health economic | Cost<sup>23</sup> | Child<br>Parent | Total costs | Baseline<br>3 months<br>6 months |
| Secondary |  |  |  |  |
| Clinical | Children's Sleep Habits Questionnaire | Child | Total score | Baseline<br>6 months |
| Clinical | Seizure-free period | Child | Time to first seizure from randomisation (days) | Randomisation<br>3 months<br>6 months |
| Clinical | Seizure remission | Child | Time to 6-months seizure remission from randomisation (days) | Randomisation<br>3 months<br>6 months |
| Clinical | Knowledge about Sleep in Childhood | Parent | Total score | Baseline<br>3 months |
| Clinical | Hospital Anxiety and Depression Scale | Parent | Total score | Baseline<br>3 months<br>6 months |
| Clinical | Insomnia Severity Index | Parent | Total score | Baseline<br>3 months<br>6 months |

<sup>1</sup> Children: CHU-9D, parents: EQ-5D, ISI
<sup>2</sup> **Sources for cost calculation (NHS and Personal Social Services):** At randomisation, 3- and 6-months: Resource use questionnaire, Case Report Form data (concomitant medication, Serious Adverse Events); At end of trial: Patient Level Information and Costing System data, Hospital Episode Statistics
<sup>3</sup> **Sources for cost calculation (Indirect and direct non-medical):** At randomisation, 3- and 6-months: Case Report Form (school absences). At baseline, 3- and 6-months: School absences, parental lost productivity [time off work], out of pocket expenses.

| Outcome type | Specific measurement variable | Collected for | Participant-level analysis metric | Measurement time-point(s) |
| --- | --- | --- | --- | --- |
| Clinical | • SleepSuite: Animal task<br>• SleepSuite: Bubble task<br>○ Shape detection<br>○ Emotion detection<br>○ Gender detection<br>• SleepSuite: Maze task | Child | Executive function (accuracy)<br>Reaction time (ms)<br>Variability (Co-efficient of variation [CV])<br>Specifically:<br>• Number of correctly named animals (count)<br>• Number of correctly solved mazes (count)<br>• Incidence rate ratio<br>• Mean reaction time (ms)<br>• Accuracy (Misses, False Alarms)<br>• Intra-individual CV | Baseline<br>3 months |
| Clinical | Health-Related Quality Of Life Measure for Children with Epilepsy | Child | Total score | Baseline<br>6 months |
| Clinical | World Health Organisation – Five Well-Being Index | Parent | Total score | Baseline<br>6 months |
| Clinical | Strengths and Difficulties Questionnaire | Child | Total score | Baseline<br>3 months<br>6 months |
| Clinical | Parenting Self Agency Measure | Parent | Total score | Baseline<br>3 months<br>6 months |
| Clinical | Sickness-related school absences | Child | Total number of days | Baseline<br>3 months<br>6 months |
| Clinical | Actigraphy | Child<br>Parent | Total sleep time (minutes)<br>(2-week average) | Baseline<br>3 months |
| Clinical | Actigraphy | Child<br>Parent | Sleep latency (minutes)<br>(2-week average) | Baseline<br>3 months |
| Clinical | Actigraphy | Child<br>Parent | Sleep efficiency (% asleep of sleep period)<br>(2-week average) | Baseline<br>3 months |
| Qualitative | Trial experience<sup>4</sup> | Child<br>Parent | Interview transcript<br>Activity booklet transcript/photos | 3 months + 3 weeks<br>6 months + 3 weeks |
| Process evaluation<br>(context, implementation, and mechanisms of impact) | • COSI system:<br>○ e-analytics<br>○ evaluation module<br>• Qualitative trial component | Parent<br>Child<br>Parent | • Module completion (%)<br>• Time spent (minutes)<br>• Intervention experience<br>○ Ease of use (Likert scale)<br>○ Techniques used (binary)<br>○ Technique helpfulness (Likert scale)<br>○ Thoughts and feelings (Likert scale)<br>• Interview transcript<br>• Activity booklet transcript/photos | Randomisation to 3 months<br>3 months + 3 weeks<br>6 months + 3 weeks |

## METHODS AND ANALYSES

### Trial design

CASTLE Sleep-E is a UK-based, multi-centre, open-label, active concurrent control, randomised (1:1), parallel-group, pragmatic superiority trial. Compared are clinical- and cost-effectiveness of standard care (SC) alone and SC augmented with a novel, tailored, parent-led CASTLE Online Sleep Intervention (SC + COSI) in reducing sleep problems in children (5 to <13 years) with RE at 3- and 6 months after randomisation. Parents and children (≥ 7years) can opt into qualitative interviews and activities to share their experiences and perceptions within 3 weeks of completion of other data collection at 3- and 6 months after randomisation. Jointly, the qualitative trial component and the COSI system (e-analytics and evaluation module) will provide information for a process evaluation (context, implementation, and mechanisms of impact).^28^

### Patient and Public Involvement

The CASTLE programme (which subsumes CASTLE Sleep-E) recruited a dedicated Patient and Public Involvement (PPI) Advisory Panel (AP) through social media and epilepsy charities in 2017. The CASTLE Advisory Panel (CAP) consists of 17 adults with experience of childhood epilepsy and five children with epilepsy (aged 6–15 years). CAP has been involved in the research programme from the funding application onward (2 CAP members are co-applicants). Full PPI details are provided in GRIPP2 Short Form in Table 2.

**Table 2.**
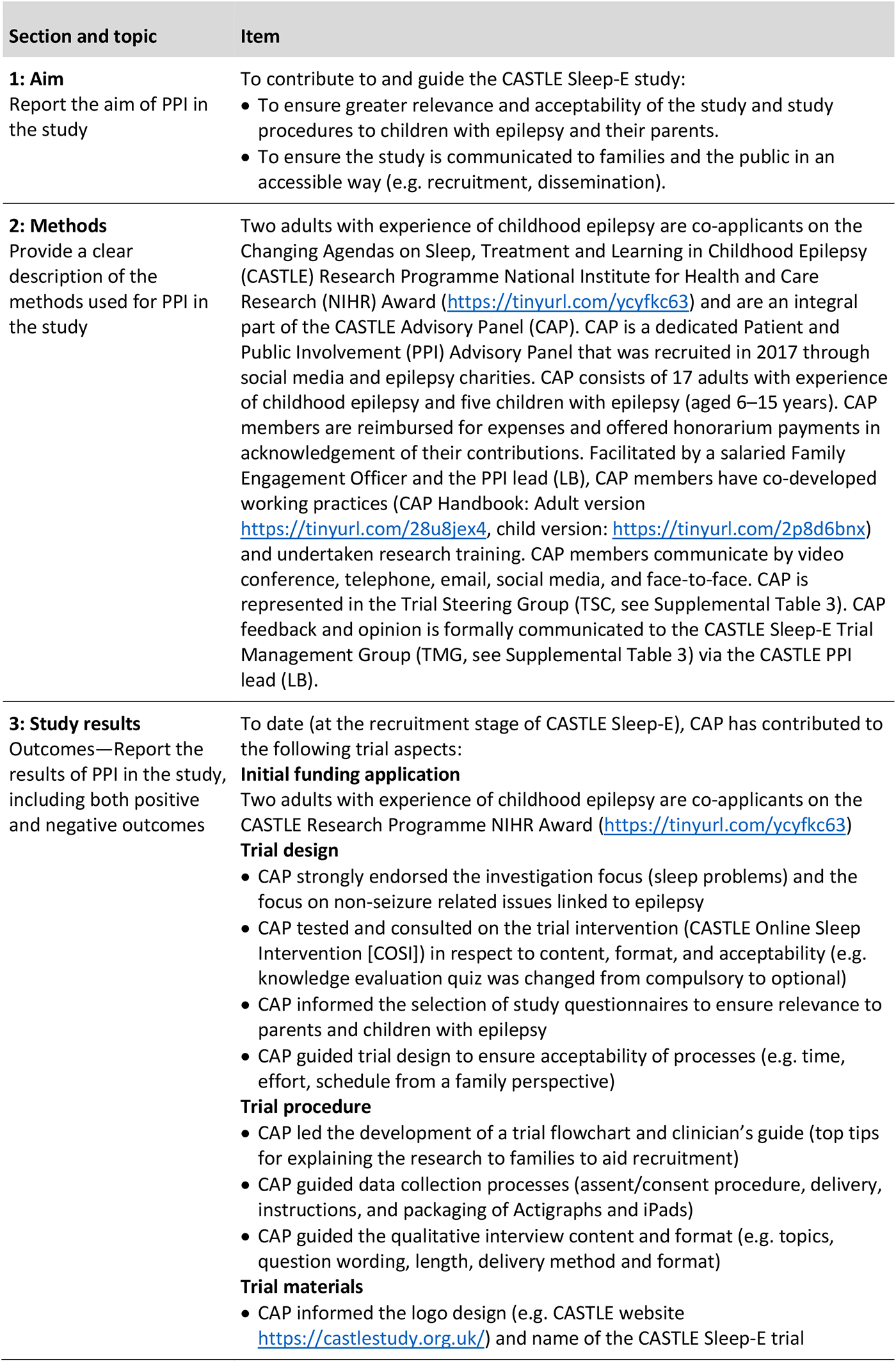
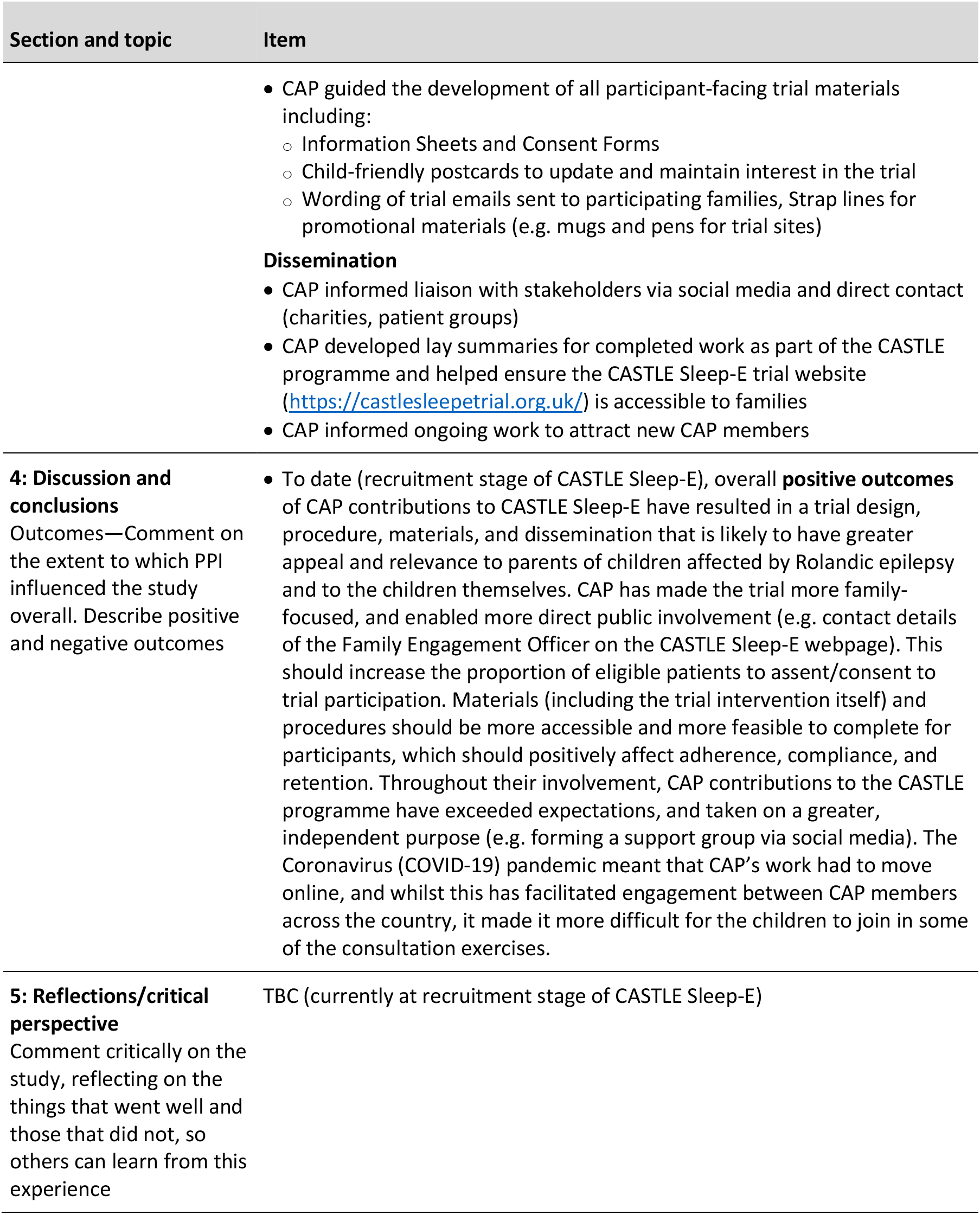
GRIPP2-Short Form (SF)21: Guidance for Reporting Involvement of Patients and the Public in research.

### Trial setting and eligibility criteria

Participants will be identified by staff in NHS out-patient general paediatric and paediatric epilepsy clinics in the UK (pre-dominantly urban setting). Eligibility criteria for trial sites include a Capacity and Capability assessment as advised for NHS site set-up by the UK Health Research Authority (HRA). The expected number of trial sites is 40 (England: 34, Scotland: 4, Wales: 1, Northern Ireland: 1). A list of trial sites can be obtained from the Trial Manager (see Supplemental Table 1).

### Intervention

Participants will be allocated to trial arms (SC or SC + COSI) using minimisation (1:1 ratio). On allocation to SC + COSI, participants will receive an email with access details to COSI. COSI consists of a self-paced, novel, tailored, e-learning package for parents of children with epilepsy that incorporates evidence-based behavioural components. Table 3 provides a brief overview; detailed reports on the development, content, and evaluation of COSI have been published.^15 16^ COSI is divided into 13 modules (1 screening for child-specific sleep problems to allow tailoring, 10 content, 1 additional resources, 1 initially hidden evaluation), of which three are compulsory (1 screening, 2 content). The non-compulsory modules are recommended based on screening outcome, but all modules are accessible, repeatable, and printable. The advice in COSI supports parents to implement general prevention techniques (e.g. good sleep hygiene) and specific behavioural change techniques (e.g. bedtime fading) relevant to their child’s sleep problems. Three months after first being given access to COSI, parents will be asked by email to complete a COSI evaluation module. At the end of a participant’s trial timeline (6 months), access to COSI will be revoked. After the trial, all families (irrespective of trial allocation) have the option to receive the COSI content in electronic format via email.

**Table 3.**
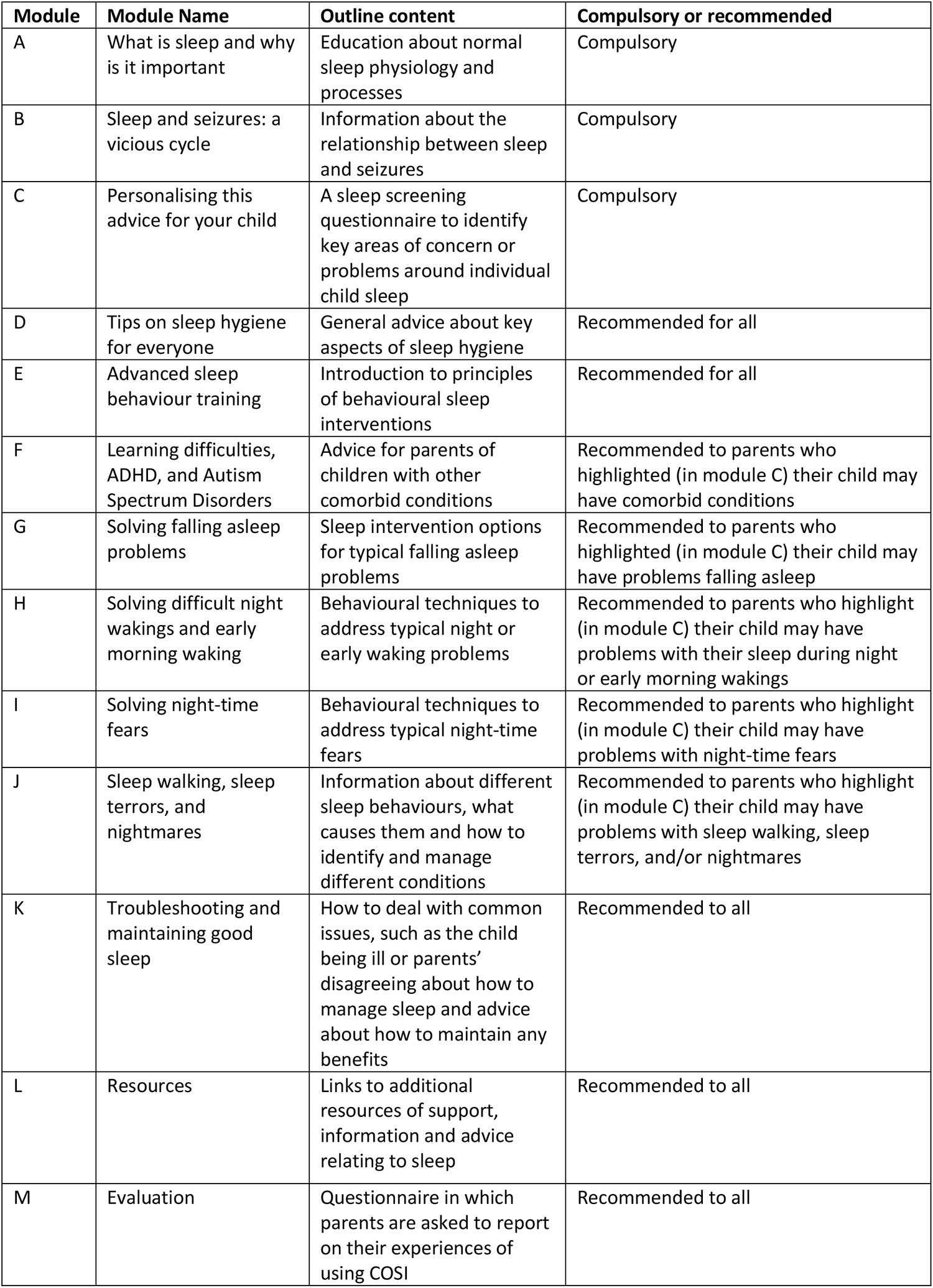
Content of the CASTLE Online Sleep Intervention (COSI)

### Fidelity, adherence, retention and acceptability

Fidelity (intervention delivery) will be monitored through e-analytics embedded in the COSI system (modules accessed, and time spent per module). Strategies to improve completion of COSI training in case of non-access include: (1) an automated text-reminder after two days; (2) an email reminder after four days; (3) a phone call from researchers who developed COSI (the Sleep Team) after six days. To improve adherence to the intervention, (1) all participants will receive a phone call from the Sleep Team six weeks after account creation; and (2) children will receive postcards with child-oriented activities (e.g. maze) at three time-points to welcome them to the trial (weeks 1–2), to stay in touch (weeks: 4–5), and to thank them for participating (weeks 4–8 post-trial). To encourage completion of the intervention evaluation, participants will receive: (1) an automated text-reminder after three days of non-completion, (2) and a phone call from the Sleep Team after eight days of non-completion. Fidelity (intervention implementation, acceptability, perceived helpfulness) will be captured jointly by the COSI evaluation module and the qualitative trial component.

### Discontinuation, withdrawal, concomitant care or interventions

Participants may discontinue the trial intervention or withdraw from the trial if (1) the parent/child withdraws consent/assent respectively; or (2) a change in the child’s condition justifies discontinuation of treatment in their clinician’s opinion. Trial site staff will record withdrawal with reason where provided in electronic Case Report Forms (eCRFs). Pseudo-anonymised data up to the time of consent withdrawal will be included in analyses in accordance with General Data Protection Regulation (GDPR)^29^ under the UK Data Protection Act 2018^30^ — the trial Data Controller relies on the legal bases of ‘public interest’ and ‘research purposes’.

To avoid confounding and to minimise participant burden, co-enrolment into other clinical trials is discouraged. Where recruitment into another trial is considered appropriate, the trial coordinating centre will discuss enrolment with the Chief Investigator (CI). Participation in the Rolandic Epilepsy Genomewide Association International Study (REGAIN: https://childhoodepilepsy.org/research-studies/regain/) is complementary (same CI).

### Outcomes and participant timeline

Outcomes are reported in Table 1 and were chosen collaboratively by children and young people with epilepsy and their parents, sleep- and epilepsy experts^8 17^ in accordance with Core Outcome Measures in Effectiveness Trials (COMET) guidelines.^31^ Psychometric properties and clinical relevance of outcomes are reported in Supplemental Table 2. Each participant will be followed up for 6 months. The participant timeline is shown in Table 4.

**Table 4.**
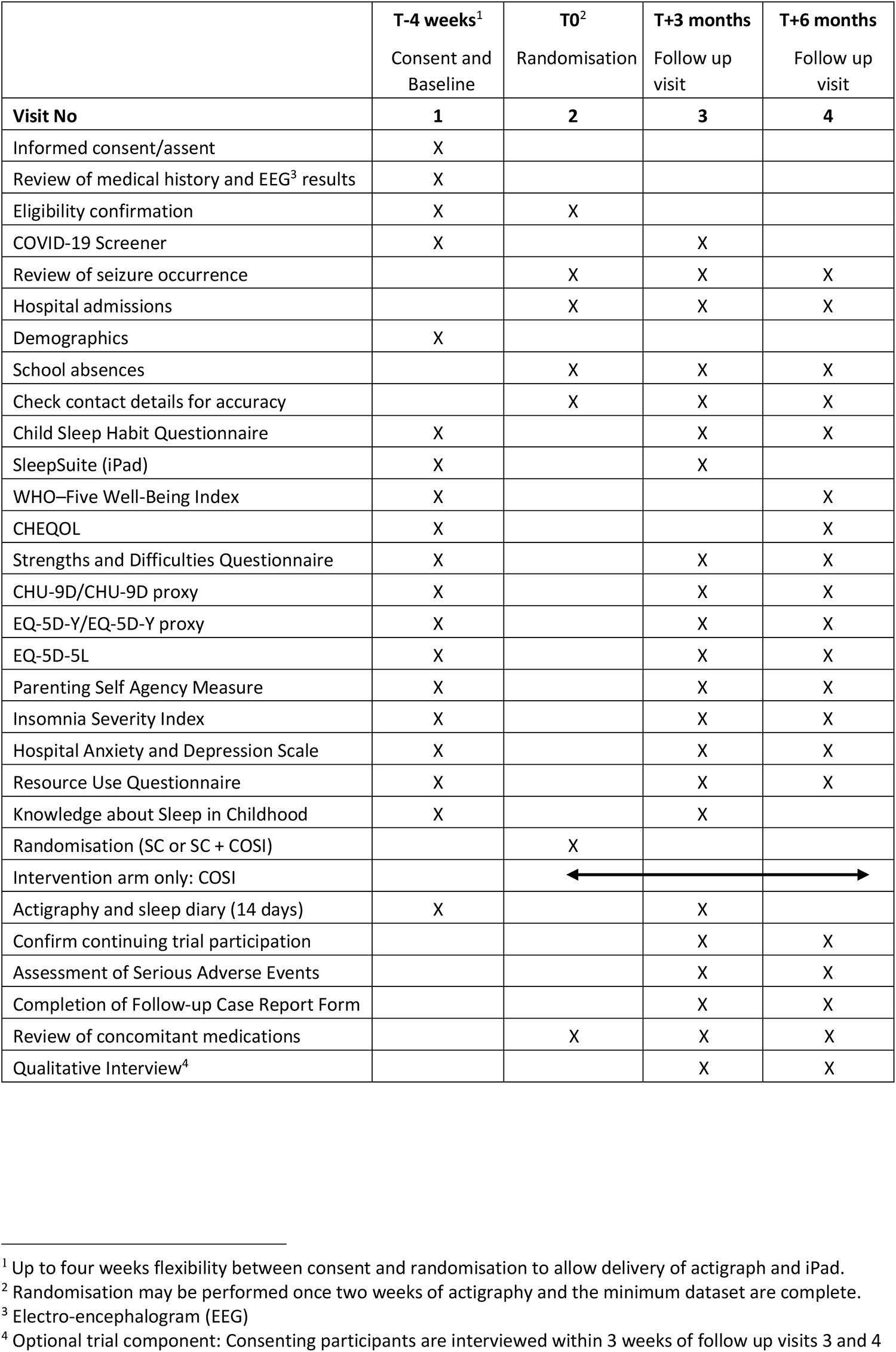
CASTLE SLEEP-E participant timeline.

### Sample size

The target sample size (110 children with RE, 55 per trial arm) was calculated based on achieving 90 % power to detect the minimal clinically important difference (MCID) in the primary clinical outcome (CSHQ) at 3 months after randomisation, accounting for 10 % expected attrition (non-parametric test with two-sided 5% significance level). MCID was defined based on an individual-focused anchor-based method,^32^ that is, ‘the smallest difference in outcome that patients perceive as beneficial and which mandates a change in patient management’.^33^ The MCID value was based on the estimated reduction in total CSHQ score required for children with epilepsy (*M* = 48.25, *SD* = 8.91)^7^ to fall at or below the diagnostic cut-off score of 41 for sleep disorders in paediatric populations.^34^

### Recruitment, stopping guidelines, interim analyses

An Independent Data and Safety Monitoring Committee (IDSMC) will monitor recruitment and make recommendations to the Trial Steering Committee (TSC) concerning trial continuation, adjustments of recruitment methods, and follow-up optimisation (see Supplemental Table 3). A traffic light approach will determine trial continuation: (1) Green: Continue if at least 30 trial sites have opened and 22 participants have been randomised by end of month 6; (2) Amber: Implement additional recruitment strategies if 15–21 participants have been randomised by end of month 6; (3) Red: If recruitment is <15 participants by end of month 6, then stopping the trial early will be discussed with the TSC. Formal interim analyses of the accumulating data will not be performed.

### Treatment allocation

Participants will be allocated with a 1:1 ratio to either SC or SC + COSI based on a computer generated adaptive restricted randomisation procedure that minimises differences between trial arms in variables likely to affect outcomes. The allocation concealment mechanism is an online, central randomisation service implemented and maintained by the Liverpool Clinical Trial Centre (LCTC). The service will be accessed within four weeks of participant enrolment (once consent and eligibility confirmed, Participant ID issued, baseline dataset completed) by trained, authorised staff at trial sites. Randomisation will trigger allocation emails to the Trial Manager at LCTC and to the relevant trial site as well as enable COSI access for participants allocated to the intervention arm. Trial sites will notify the participant’s General Practitioner (GP) of the treatment allocation by letter (electronic or hard copy, depending on preference).

### Blinding

Only quantitative data analysts will be blinded (Participant IDs do not reveal treatment allocation). All other stakeholders (participants, parents, healthcare providers, data collectors, qualitative researchers) will be aware of the allocated intervention. Emergency unblinding procedures are therefore unnecessary.

### Assent and consent

Potentially eligible children will be screened at trial centres by trained site staff. Screening outcome will be documented. Eligible children with interested parents will be invited to participate and provided with a Patient Information Sheet and Consent Form electronically and/or hard-copy (PISC, three versions: Parent, child [5–6 and 7–12 years]). Sufficient time will be allowed for discussion of the trial and the decision to assent/consent to trial entry and the optional qualitative component. Assent (children aged 7-12 years) and Consent (parents) may be given face-to-face or remotely and will be electronically captured in a secure Consent Database managed by LCTC. Reasons for declining participation will be asked, but it will be made clear that children and parents do not have to provide a reason.

### Data collection and management

Data collection will be carried out electronically except for Serious Adverse Events and Participant Transfer Forms (hard copy). At consent/assent, site staff will enter patient medical history (including electro-encephalogram), eligibility confirmation, COVID-19 screening, and demographics (see Table 4) into eCRFs stored in a secure Data Management System managed by LCTC. Trial participation will be added to the patient’s medical records alongside their unique Participant ID.

Consent- and Contacts Databases are securely linked. The addition of a new participant will trigger email notifications to the Parents containing access links to baseline assessments (see Table 4) and the Sleep Team who will access the Contacts Database to arrange the delivery of an iPad pre-configured by LCTC (optionally fitted with pre-paid SIMs) and two actigraphs with supporting documents. iPads (Generations 7–8, iOS 15.2 or 15.3) will be used to access the SleepSuite App, (V 1.4)^35^, which assesses executive functions in child-friendly, interactive games (e.g. popping virtual balloons with smiling children’s faces). Access requires the Participant ID and is only possible at pre-specified trial time-points (see Table 4). Data is only stored on the iPad until the test-session completion, then automatically uploaded to a cloud-based server, and then securely downloaded for analyses by authorised LCTC staff. Families lacking other means of internet access can use iPads fitted with pre-paid SIMS to access other online trial materials (including email).

Actigraphs (Micro Motionlogger® Watch and Watchware Software V 1.99.17.4, Ambulatory Monitoring, Inc., NY: USA) will be used to collect 14 days of objective sleep data from child and parent. Concurrent sleep diaries (hard copies) will be completed by the parent with or without child input. At the end of the baseline period, actigraphs will be returned to the Sleep Team via pre-paid courier, and iPads to LCTC via trial sites. The Sleep Team will download and securely store pseudo-anonymised (using Participant IDs) actigraphy data for pre-processing (manual selection of sleep periods cross-checked against sleep diaries) per night at participant-level. Summary variables (sleep latency, total sleep time and sleep efficiency) are then automatically calculated by actigraph software, manually collated and securely transferred electronically to LCTC for trial-level analyses by the Trial Statistician.

Participants will be randomised to trial arms during a telephone/video call or clinic visit only *after* site staff have confirmed that baseline data (see Table 4) is complete, and eligibility, consent/assent, and contact details are still valid. Data collection will be repeated 3- and 6 months after randomisation (see Table 4).

The Qualitative Research Team will access the Contacts Database to schedule audio-recorded interviews with children and parents who consented/assented to this optional trial component. Interviews (audio- or audio-video) will take place remotely within 3 weeks of completion of other data collection at 3- and 6 months after randomisation. Parents and children will be interviewed together or separately as preferred. Parents and children will have the opportunity to think through their ideas prior to the interview (as proposed by parents and children from the CASTLE Advisory Panel). Children will be invited to complete activity booklets in advance of their interviews (the booklets will be mailed or e-mailed one week prior to their interview); the content they complete will support the interview. Parents will receive a list of proposed questions/topics. Children will be able to share the booklet with the Qualitative Team (e.g. screen or photograph sharing, verbal description).

The direct costs of health and personal social services, and indirect costs of productivity losses and school absenteeism will be collected using a Resource Use Questionnaire administered at baseline and during follow-up visits. Other data such as concomitant medications, study visits and Adverse Events will be collected using eCRFs. Trial participants’ use of secondary care services will be collected from Patient-Level Information and Costing Systems (PLICS) data obtained from the finance departments of each recruiting hospital; and based on Hospital Episode Statistics (HES) obtained from NHS Digital at the end of the trial. PLICS and HES data will be pseudo-anonymised and transferred securely to the trial health economists at Bangor University.

### Data quality, security, and trial oversight

Reliability, validity, and clinical relevance of outcomes are reported in Supplemental Table 2. Processes to promote quality and security of collected data include general local training of site staff and research teams (Good Clinical Practice); and trial-specific training in the use of electronic forms and databases by LCTC. LCTC will request to see evidence of appropriate training and experience of all trial staff. Staff will be signed off as appropriately qualified by the CI. Electronic data capture provides several in-built validity and security checks (e.g. data type, range, and missingness checks in eCRFs, SleepSuite use/access restrictions). Some electronic and all hard-copy data will be repeat checked (e.g. eligibility, contact details). Data processing requiring more subjective judgement will be performed by minimum of two trained researchers on at least a subset of data (i.e. manually-assisted selection of actigraphy sleep period; thematic and content analysis of qualitative data).

Data will be processed and stored in accordance with GDPR under the UK Data Protection Act 2018. Central data monitoring will be performed by LCTC who will raise and resolve queries with site and research teams within the online system. The University of Liverpool is registered with the Information Commissioners Office. LCTC will receive trial participants’ HES identifiers for secure transfer to the Health Economic team, who will access, securely store, and dispose of HES data in accordance with the Bangor University and NHS Digital Data Sharing Framework Contract.

### Statistical methods

Statistical analyses of all but health economic and qualitative data will be performed by the Trial Statistician (LCTC) using SAS software, Version 9.4 or later. Intention-To-Treat (ITT) will be the main analysis strategy for primary and secondary outcomes (see Table 1 and Table 5). Statistical significance will be set at the conventional two-sided 5 % level; clinical relevance will be based on previous research (see Supplemental Table 2). Point estimates with 95 % two-sided confidence intervals will be reported adjusted and unadjusted for covariates. No multiplicity adjustments will be made (only one primary clinical outcome, uncorrected secondary outcome analyses).

**Table 5.** Analysis plan for outcome variables in CASTLE Sleep-E. Further analyses details are reported in-text.

| Outcome type | Specific measurement variable | Hypothesis | Method of analysis |
| --- | --- | --- | --- |
| **Primary** |  |  |  |
| Clinical | Children's Sleep Habits Questionnaire | Total score lower in intervention arm at 3 months | Linear mixed effect regression<br>• Fixed effects:<br>○ Intervention (binary)<br>• Random effects:<br>○ Trial site (categorical)<br>• Co-variates:<br>○ Baseline score<br>○ Use of sleep medication (binary) |
| Health economic | Cost<sup>9</sup> per quality-adjusted life year gained<sup>10</sup> | Incremental Cost Effectiveness Ratio for intervention below NHS threshold | Cost-effectiveness analysis |
| **Secondary** |  |  |  |
| Clinical | Children's Sleep Habits Questionnaire | Total score lower in intervention arm at 6 months | Linear mixed effect regression (as before) |
| Clinical | Seizure-free period | Time to first seizure (days) differs between trial arms at 3 and 6 months | Survival analyses<br>• Kaplan-Meier curves by trial arm<br>• Cox proportional hazards regression (if applicable)<br>○ Co-variates:<br>▪ Use of sleep medication (binary)<br>▪ Trial site (categorical) |
| Clinical | Time to 6-months seizure remission from randomisation (days) | Time to 6-months seizure remission (days) differs between trial arms at 6 months | Survival analyses (as before) |
| Clinical | Knowledge about Sleep in Childhood | Total score differs between trial arms at 3 months | Linear mixed effect regression (as before) |
| Clinical | Hospital Anxiety and Depression Scale | Total score lower in intervention arm at 3 and 6 months | Linear mixed effect regression (as before) |
| Clinical | Insomnia severity index | Total score lower in intervention arm at 3 and 6 months | Linear mixed effect regression (as before) |

<sup>9</sup> NHS and PSS perspective; Alternative perspective: Societal (Indirect and direct non-medical costs)
<sup>10</sup> Children: CHU-9D, parents: EQ-5D, ISI

| Outcome type | Specific measurement variable | Hypothesis | Method of analysis |
| --- | --- | --- | --- |
| Clinical | • SleepSuite: Animal task<br>• SleepSuite: Bubble task<br>○ Shape detection<br>○ Emotion detection<br>○ Gender detection<br>• SleepSuite: Maze task | Executive function, reaction time, and variability differ between trials arm at 3 months | • Poisson/zero-inflated negative binomial regression (depending on presence of overdispersion)<br>• 2 x 2 multi-variate repeated-measures Analysis of Variance (ANOVA)<br>Factors: Time (PM/AM) x Intervention (Pre/Post)<br>Fitted per detection task (Shape, Emotion, Gender)<br>• Linear mixed effect regression (as before)<br>○ Fixed effects:<br>▪ Intervention (binary)<br>▪ Level played<br>○ Co-variates:<br>▪ Baseline score |
| Clinical | Health-Related Quality Of Life Measure for Children with Epilepsy | Total score differs between trial arms at 6 months | Linear mixed effect regression (as before) |
| Clinical | World Health Organisation – Five Well-Being Index | Total score differs between trial arms at 6 months | Linear mixed effect regression (as before) |
| Clinical | Strengths and Difficulties Questionnaire | Total score differs between trial arms at 3 and 6 months | Linear mixed effect regression (as before) |
| Clinical | Parenting Self Agency Measure | Total score differs between trial arms at 3 and 6 months | Linear mixed effect regression (as before) |
| Clinical | Actigraphy: Total sleep time<sup>11</sup> (2-week average) | Total score differs between trial arms at 3 months | Linear mixed effect regression (as before) |
| Clinical | Actigraphy: Sleep latency (2-week average) | Total score differs between trial arms at 3 months | Linear mixed effect regression (as before) |
| Clinical | Actigraphy: Sleep efficiency (2-week average) | Total score differs between trial arms at 3 months | Linear mixed effect regression (as before) |
| Clinical | Sickness-related school absences | Total days differs between trial arms at 3 and 6 months | Poisson mixed-effects regression |

<sup>11</sup> Assessed in parent and child

| Outcome type | Specific measurement variable | Hypothesis | Method of analysis |
| --- | --- | --- | --- |
| Qualitative | Trial experience<sup>12</sup> | Not applicable (inductive) | Thematic analysis (interpretive, reflexive, and conceptual analytical approach)<br>• Discrete sets:<br>○ Intervention/Control<br>○ Child/parent<br>○ Engagement with intervention/lack thereof<br>○ Decision making types<br>○ Responses/experiences<br>• First separately for child and parent<br>• Secondly jointly (dyad)<br>• Comparisons to selective objective data as emerging from analysis, including, but not limited to:<br>○ Anxiety measures<br>Actigraphy-based sleep outcomes |
| Process evaluation (context, implementation, and mechanisms of impact). | • COSI system:<br>○ e-analytics<br>○ evaluation module<br>• Qualitative trial component: See previous entry | n/a | Descriptive only.<br>No formal statistical testing will be undertaken.<br>• The number and percentage of participants who accessed, attempted, and completed the quizzes for overall and for each module<br>• The completion rate of COSI will be derived for each participant as the number of questions the participant attempted out of the total number of questions available within each quiz for each module.<br>• An overall completion rate of COSI will be derived as a percentage of the questions attempted out of the total number of questions available in all the quizzes combined. The mean and standard deviation of this percentage will be presented as well as the median and range (minimum and maximum). |

<sup>12</sup> **Source data for trial experience:** Qualitative interviews (parents and children individually and as dyad), activity booklets (children only)

Sensitivity analyses will be carried out if the amount of missing data is greater than 10 %. Multiple imputation will be used to assess the robustness of the analysis to missing primary outcome data. The multiple imputation method will follow published guidelines.^36^ PROC MI in SAS will be used to generate 50 complete data sets. The imputation model will include all variables included in the primary outcome analysis model. The overall summary adjusted mean difference will be presented with 95 % confidence intervals, to assess the sensitivity of the primary analysis to missing data. All analyses will be reported in accordance with the Consolidated Standards of Reporting Trials Checklist (CONSORT)^37^ and regardless of statistical significance.

### Health economic evaluation

The economic analysis will be performed by the trial health economists at Bangor University. The primary analysis will adopt an NHS and Personal Social Services perspective and, based on Quality-Adjusted Life Years (QALYs) as a measure of health outcome, estimate the incremental cost-effectiveness ratio from an incremental analysis of the mean costs and QALYs for the intervention and control trial arms.^38^ Data assumed to be missing at random will be imputed using multiple imputation by chained equations.^39^

Sensitivity analyses will be conducted to test whether, and to what extent, the incremental cost effectiveness ratio is sensitive to key assumptions in the analysis (e.g. unit prices, different utility estimates CHU-9D vs. EQ-5D-Y). The joint uncertainty in costs and QALYs will be addressed through application of bootstrapping and estimation of cost-effectiveness acceptability curves.^40^ Alternative scenarios considering a broader cost perspective (including indirect costs, such as school absences and loss of productivity, valued by reference to published sources), and a range of outcomes (including parental QALYs, measured using the EQ-5D-5L and ISI) will be conducted. Inclusion of spillover disutility^41^ (impact on parents’ utility) will be based on the NICE reference case specification^42^ that all QALYs are of equal weight and calculated assuming additive effects. Health-economic findings will be reported according to the Consolidated Health Economic Evaluation Reporting Standards (CHEERS).^43^

### Qualitative component

Child and parent interviews will be analysed by the Qualitative Research Team using an interpretive, reflexive, and conceptual analytical approach. Audio-recordings of interviews will be transcribed and thematically analysed in discrete sets (e.g. intervention/control, child/parent, engagement/lack of engagement with intervention, types of decision-making, different responses/experiences). Parent and child transcripts will first be analysed separately, and then as dyads. All data will be used for synthesis. Thematic and content analysis will be used for child activity booklets (text and images). Qualitative- and selected quantitative data (e.g. anxiety measures, actigraphy data) will be compared, as appropriate.

### Harms

A flowchart of Adverse Event (AE) reporting requirements is shown in Supplemental Figure 1. Harms severity and causality will be graded by the investigator responsible for the care of the participant based on categories shown in Supplemental Table 4. If any doubt about causality exists, the local investigator should inform LCTC who will notify the CI. In case of discrepant views, the Research Ethics Committee (REC) will be informed of both views. Seriousness and expectedness of AEs will be defined based on International Council for Harmonisation of Technical Requirements for Pharmaceuticals for Human Use Definitions and Standards for Expedited Reporting (ICH E2A, ref: CPMP/ICH/377/95). Expectedness will be assessed by the CI. The only expected AEs in CASTLE Sleep-E are mild and transient worsening of sleep behaviours targeted by the trial intervention. Safety data will be quality-checked by a statistician not otherwise involved in the trial. Safety analysis will include all patients randomised and starting treatment and be presented descriptively split by treatment arm.

### Auditing

The CI will ensure that the trial team conducts monitoring activities of sufficient quality and quantity (e.g. protocol adherence, consent/assent, data quality). The Sponsor will delegate monitoring duties and activities to LCTC. The CI and LCTC will inform the Sponsor of any concerns. Auditing does not meet the National Institute for Health and Care Research (NIHR) or SPIRIT Statement definitions of independence^19 44^ as auditors (LCTC and CI) are part of the trial team.

### Protocol amendments

Substantive protocol amendments will be notified to HRA via the UK’s Integrated Research Application System (IRAS). Trial sites will receive an amendment pack of HRA- and REC-approved changes and unless an objection is received within 35 days, the trial will continue at site with a GO LIVE email.

### Ancillary and post-trial care

King’s College London (KCL) holds insurance against claims from participants for harm caused by their participation in this clinical study; compensation can be claimed in case of KCL negligence.

### Ethics and dissemination

The CASTLE Sleep-E protocol was approved by the HRA East Midlands – Nottingham 1 REC, reference: 21/EM/0205. Trial results will be disseminated to scientific audiences in peer-reviewed publications and conferences, and — with the help of the CASTLE Advisory Panel (parent and child experts-by-experience), relevant charities (e.g. Epilepsy Action, Epilepsy Society and Cerebra) and professional groups (e.g. Royal College of Paediatrics and Child Health, Epilepsy Specialist Nurses Association) — as plain language summaries to families, other professional groups, managers, commissioners, and policy makers. Pseudo-anonymised Individual Patient Data and associated documentation (e.g. protocol, statistical analysis plan, annotated blank Case Report Form) will be made available after dissemination on reasonable request.

### Registration details

ISRCTN registry (Trial ID: ISRCTN13202325, prospective registration 09/Sep/2021). The World Health Organisation Trial Registration Data Set (Version 1.3.1) for CASTLE Sleep-E is shown in Supplemental Table 1.

## Data Availability

Pseudo-anonymised Individual Patient Data and associated documentation (e.g. protocol, statistical analysis plan, annotated blank Case Report Form) will be made available after dissemination on reasonable request.

## Author Statement

### Contributorship

Conception of study/award holders: PG, DKP (Chief Investigators); CAP, CT-S, HH, LW, BC, CM, DH, LB (Co-Investigators)

Topic expertise (alphabetic topic order):

- Core outcome set development: CAP, LB, BC, AC, HC, PG, DH, CM, HM, DKP, MR, CT-S, PRW
- Epilepsy expert-by-experience: CAP
- Epilepsy: DKP
- Health economic evaluation: WASH, DH, EW
- Intervention development: GC, PG, HH, DKP, LW
- Patient and Public Involvement (Advisory Panel and Family Engagement): CAP, LB, BC, CM
- Responsibility for Patient-Reported Outcomes: CM
- Programme management: AC
- Qualitative research: CAP, LB, BC, HS
- Sleep: GC, PG, HH, LW
- Statistical analyses: CT-S, VW
- Trial management: NA-N, CS, LS-E

All authors contributed to the design and refinement of the study protocol. The final protocol manuscript was written by KCD, incorporated feedback from DKP, PG, CT-S, LW, BC, CM, DH, LB, GC, LS-E, CS, VW, and was approved for submission by the Chief Investigator (DKP). GRIPP2 content was checked for accuracy by LB. Sponsor name and contact information are provided in Supplemental Table 1. Details of trial committees and other groups and individuals overseeing the trial are listed in Supplemental Table 3. Trial site Principal Investigators will be listed alphabetically in resulting publications as members of the CASTLE Sleep-E Consortium in the Acknowledgements section. There has not been and will not be any use of hired writers.

### Disclaimer

To avoid potential bias, neither the funder nor the sponsor of this trial has any role in or authority over the design, execution, analyses, interpretation of data, or result dissemination.

### Patient and Public Involvement

The CASTLE Research Programme has benefitted from extensive Patient and Public Involvement (PPI) via a dedicated CASTLE Advisory Panel from its funding application onward. PPI details are reported in the Methods and Analyses section, subsection Patient and Public Involvement and in Guidance for Reporting Involvement of Patients and the Public (GRIPP2) Short Form in Table 2.

### Patient consent for publication

Not applicable.

### Provenance and peer review

Not commissioned, externally peer reviewed.

### Supplemental material

This content has been supplied by the author(s). It has not been vetted by BMJ Publishing Group Limited (BMJ) and may not have been peer-reviewed. Any opinions or recommendations discussed are solely those of the author(s) and are not endorsed by BMJ. BMJ disclaims all liability and responsibility arising from any reliance placed on the content. Where the content includes any translated material, BMJ does not warrant the accuracy and reliability of the translations (including but not limited to local regulations, clinical guidelines, terminology, drug names and drug dosages), and is not responsible for any error and/or omissions arising from translation and adaptation or otherwise.

### Licence statement

I, the Submitting Author has the right to grant and does grant on behalf of all authors of the Work (as defined in the below author licence), an exclusive licence and/or a non-exclusive licence for contributions from authors who are: i) UK Crown employees; ii) where BMJ has agreed a CC-BY licence shall apply, and/or iii) in accordance with the terms applicable for US Federal Government officers or employees acting as part of their official duties; on a worldwide, perpetual, irrevocable, royalty-free basis to BMJ Publishing Group Ltd (“BMJ”) its licensees and where the relevant Journal is co-owned by BMJ to the co-owners of the Journal, to publish the Work in BMJ Open and any other BMJ products and to exploit all rights, as set out in our licence.

The Submitting Author accepts and understands that any supply made under these terms is made by BMJ to the Submitting Author unless you are acting as an employee on behalf of your employer or a postgraduate student of an affiliated institution which is paying any applicable article publishing charge (“APC”) for Open Access articles. Where the Submitting Author wishes to make the Work available on an Open Access basis (and intends to pay the relevant APC), the terms of reuse of such Open Access shall be governed by a Creative Commons licence – details of these licences and which Creative Commons licence will apply to this Work are set out in our licence referred to above.

---

**Supplemental Table 1.**
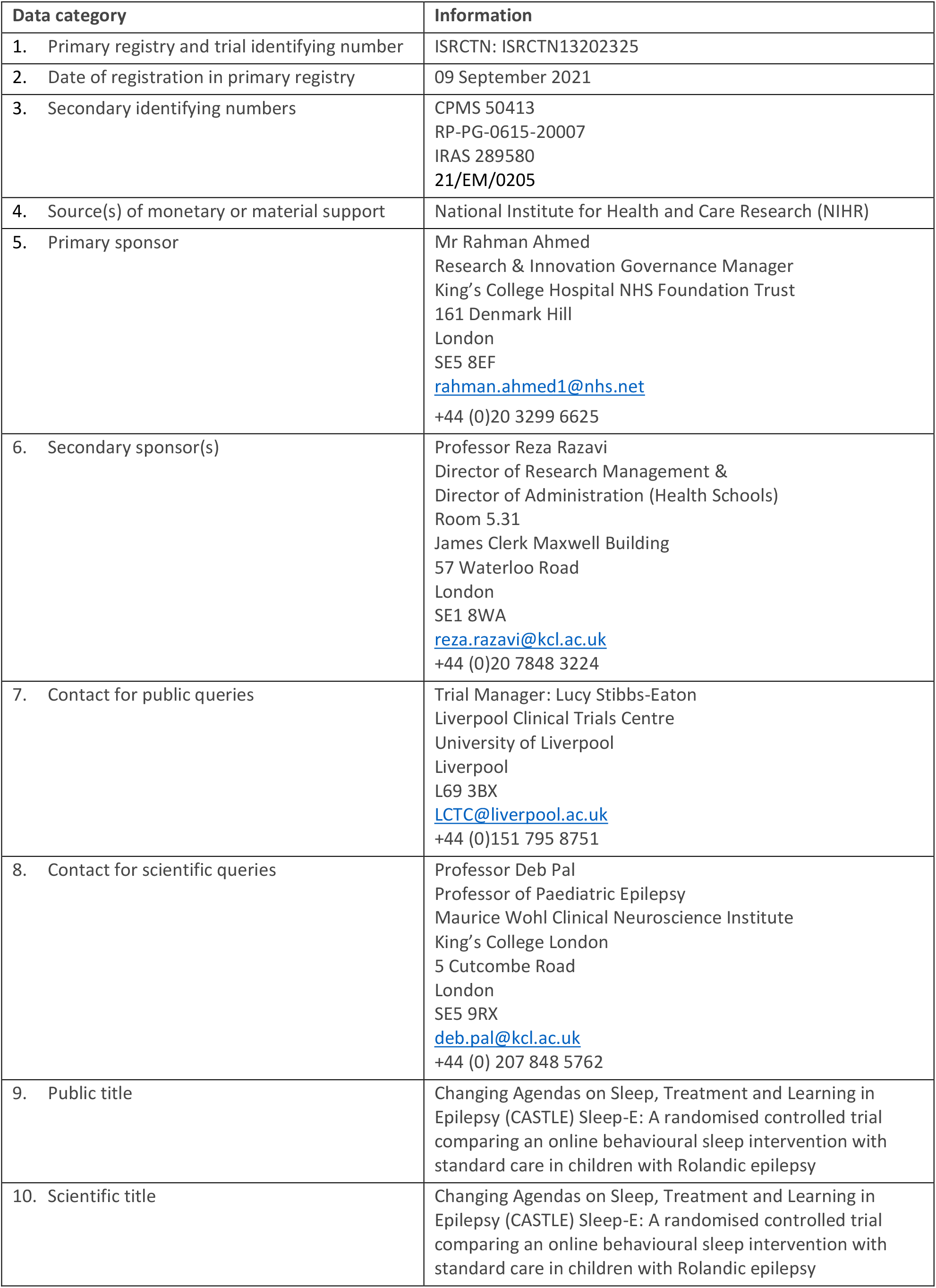
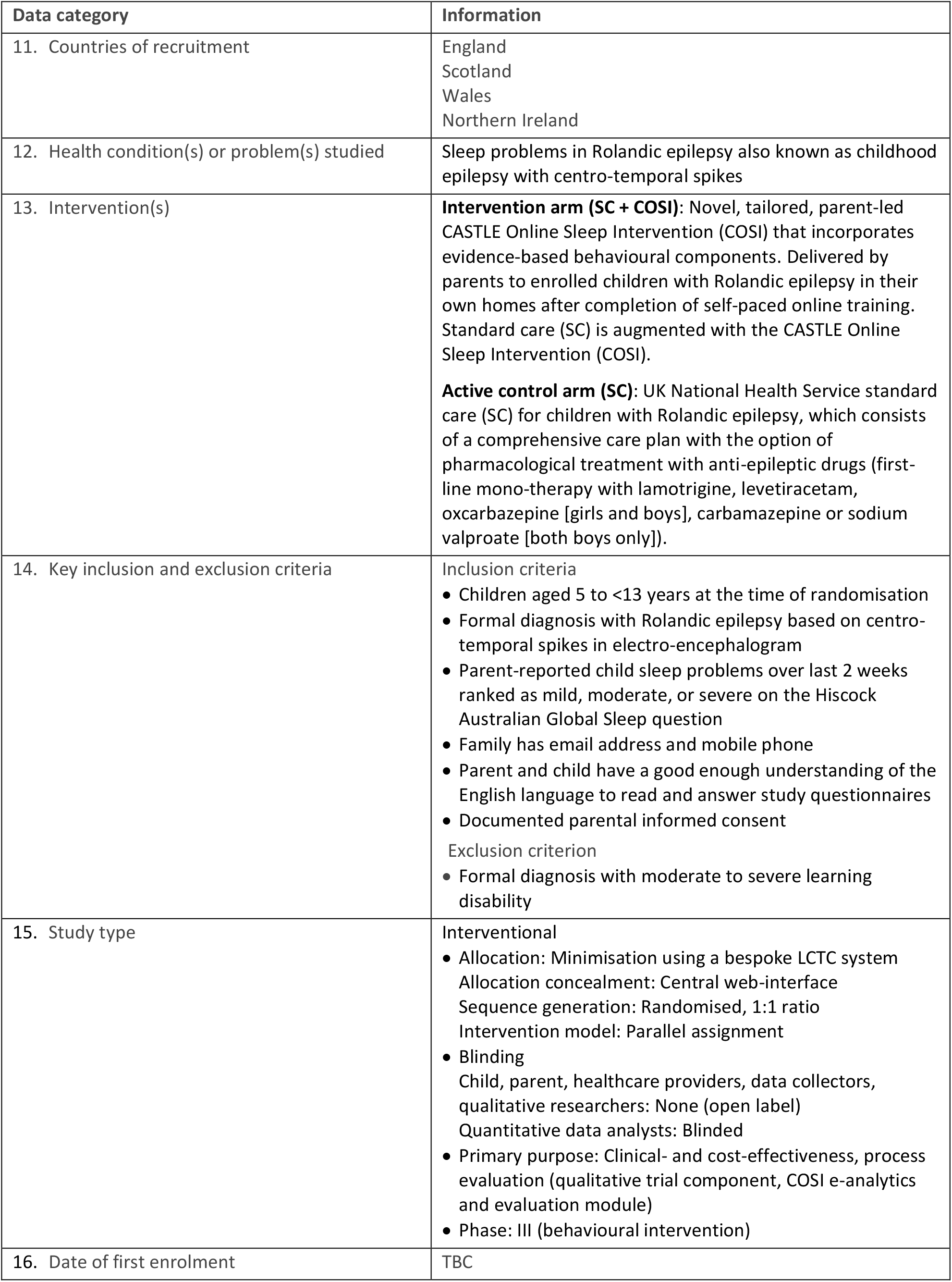
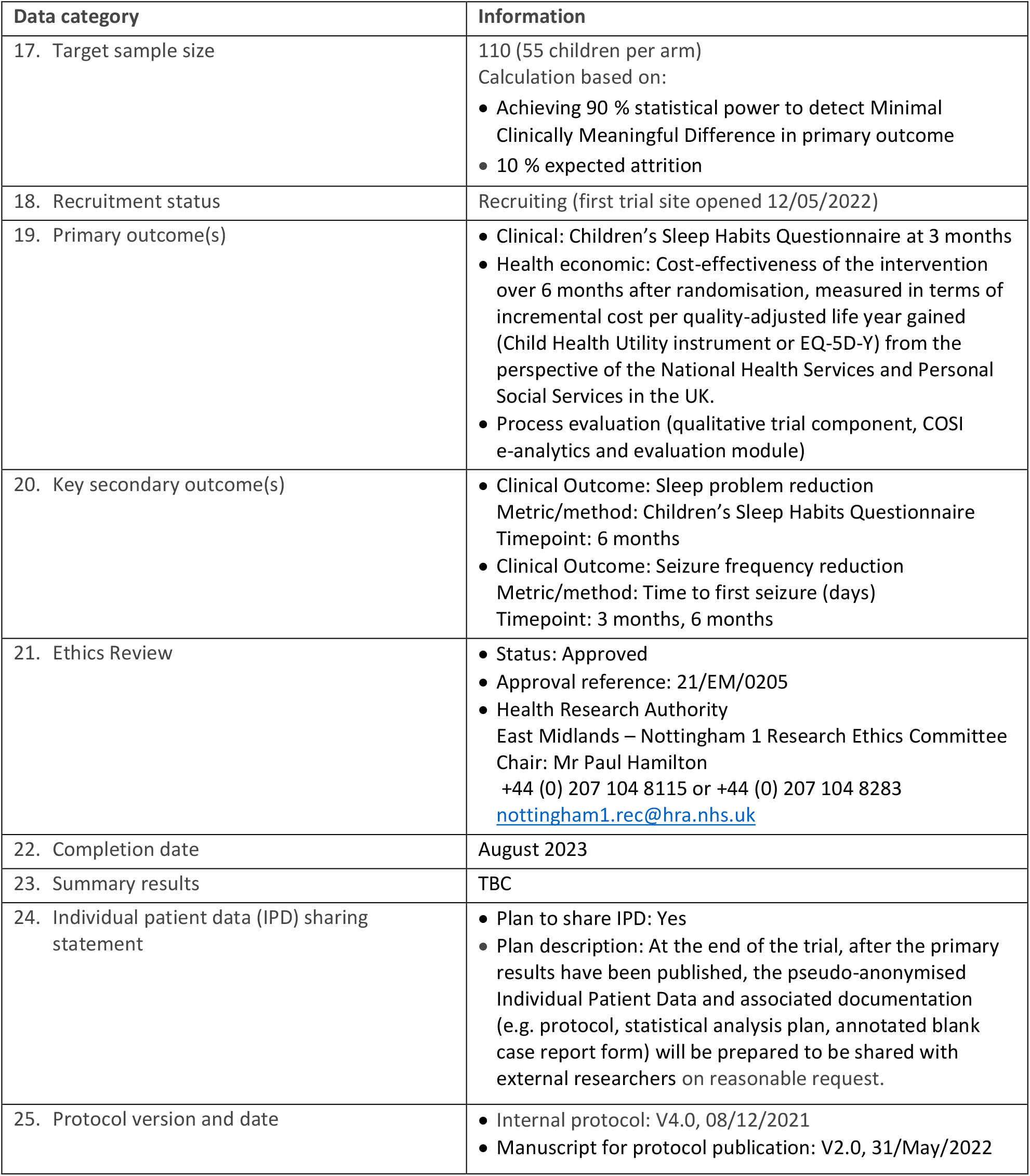
World Health Organization Trial Registration Data Set (Version 1.3.1) for CASTLE Sleep-E.

**Supplemental Table 2.**
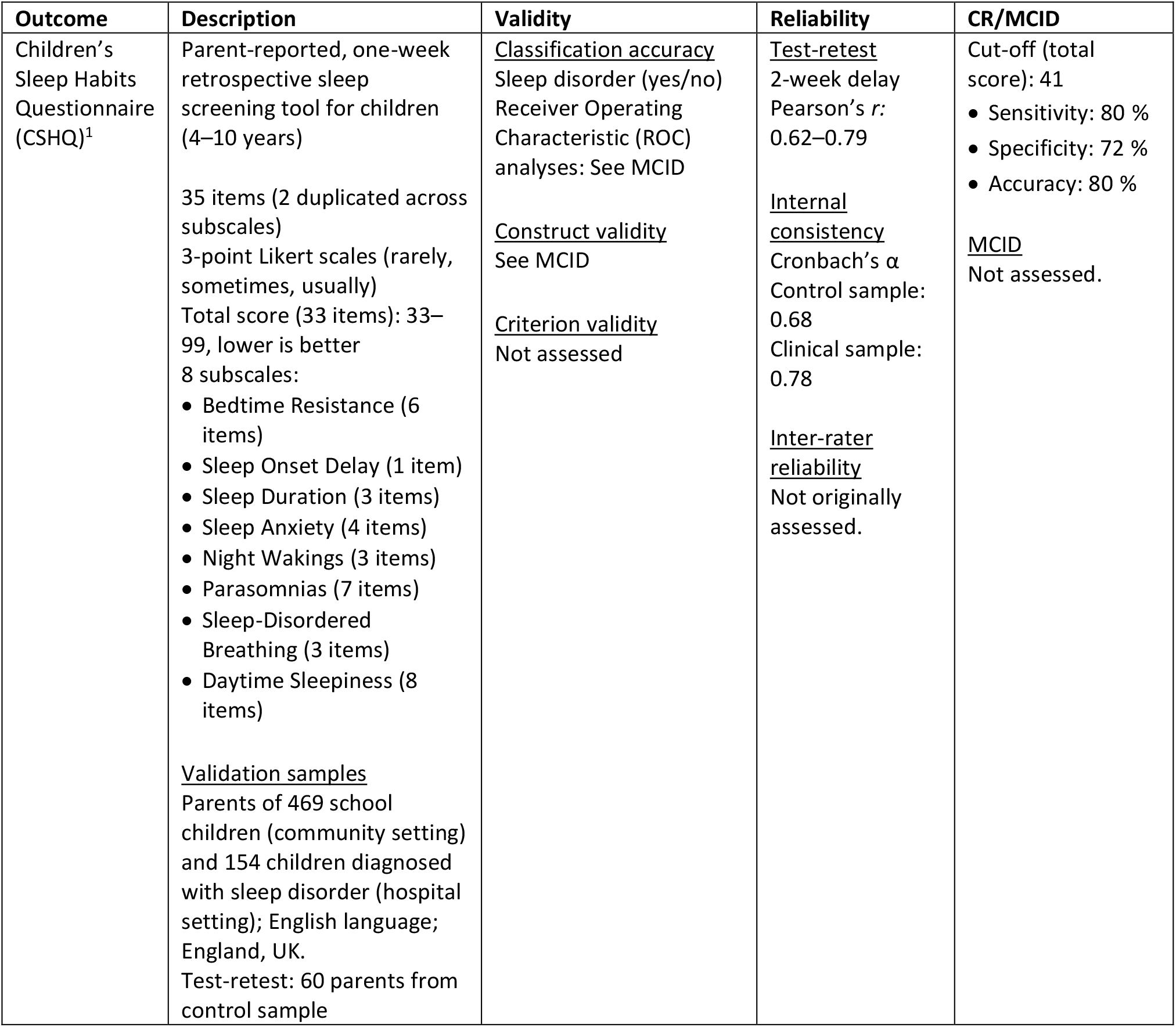
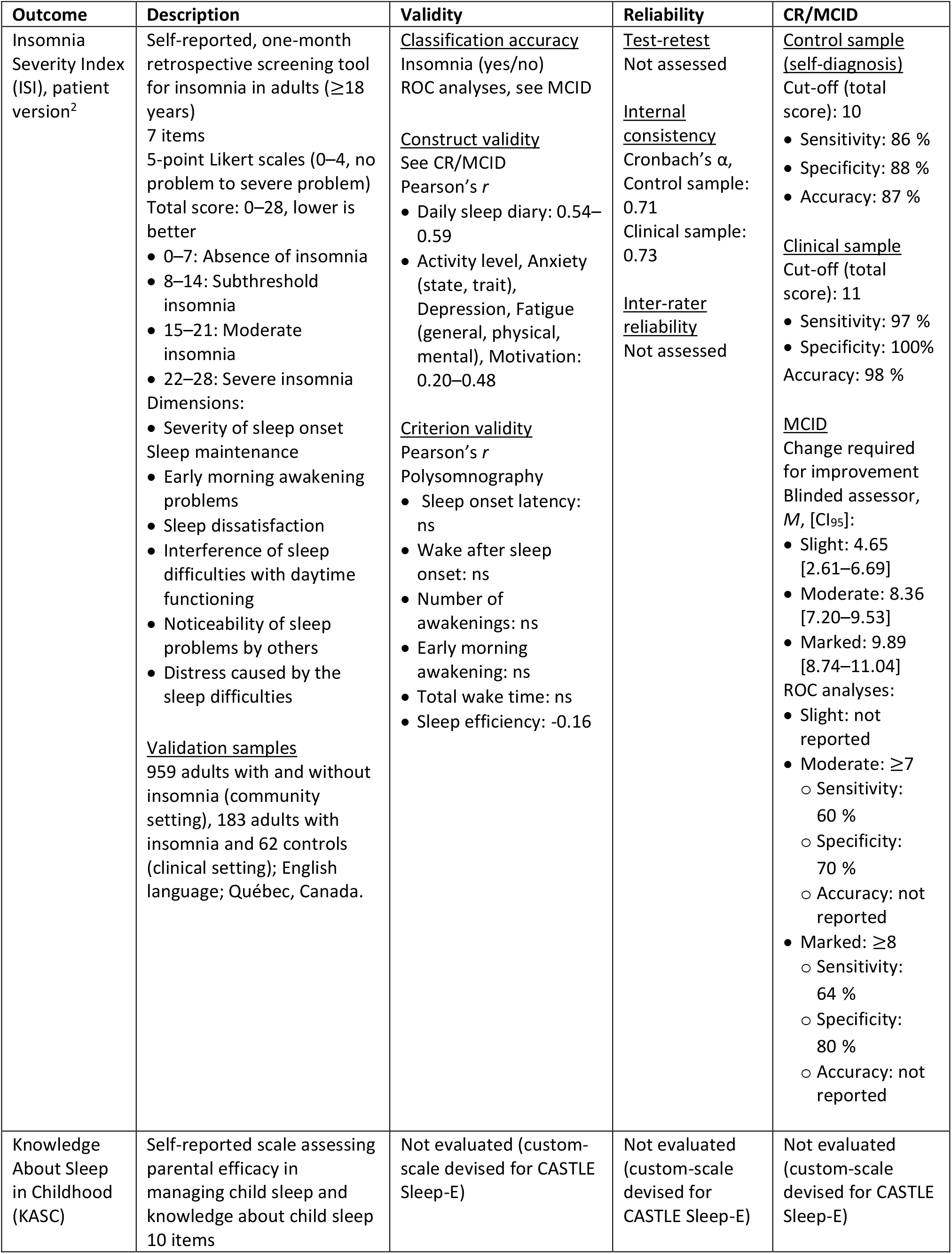
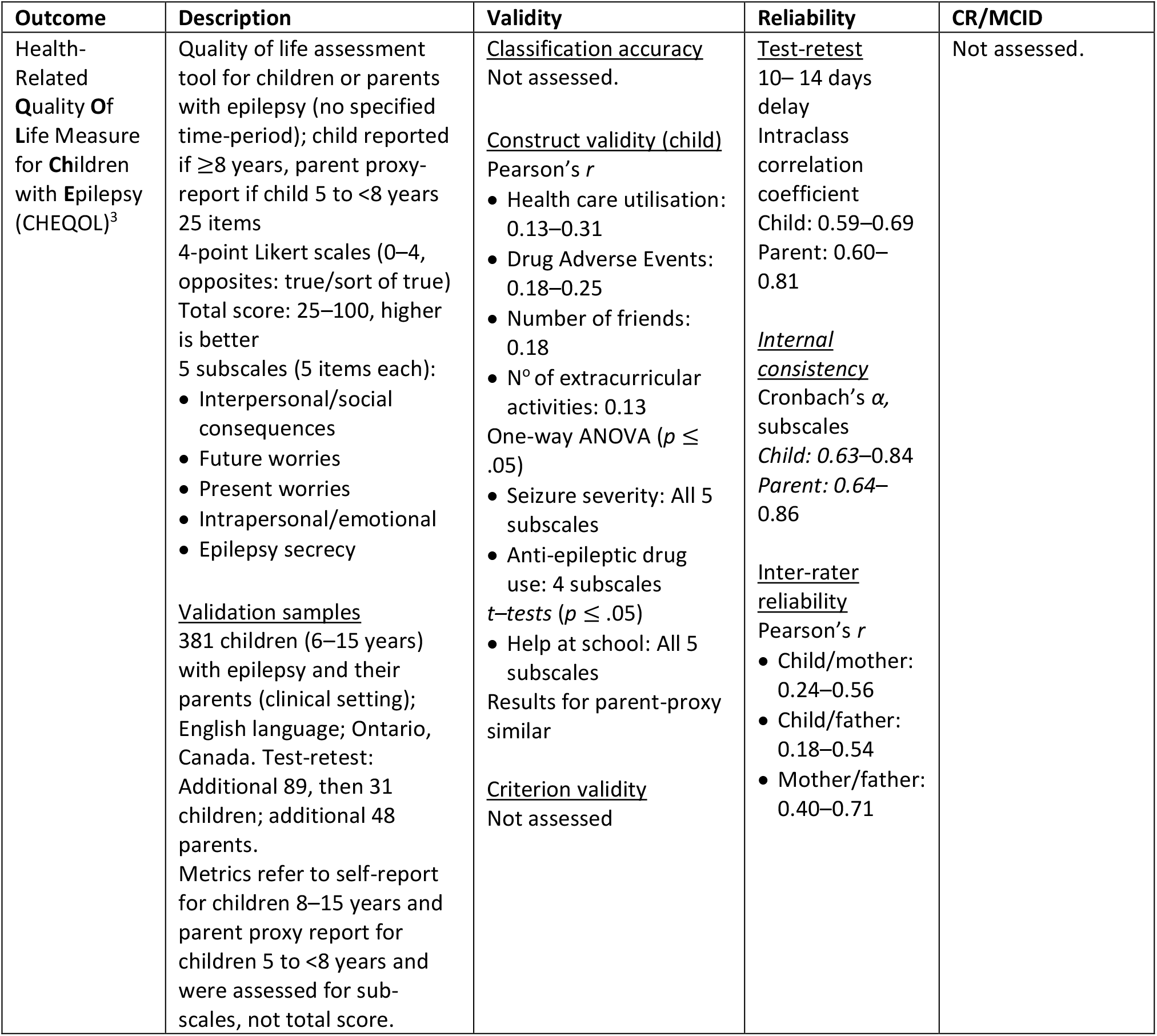
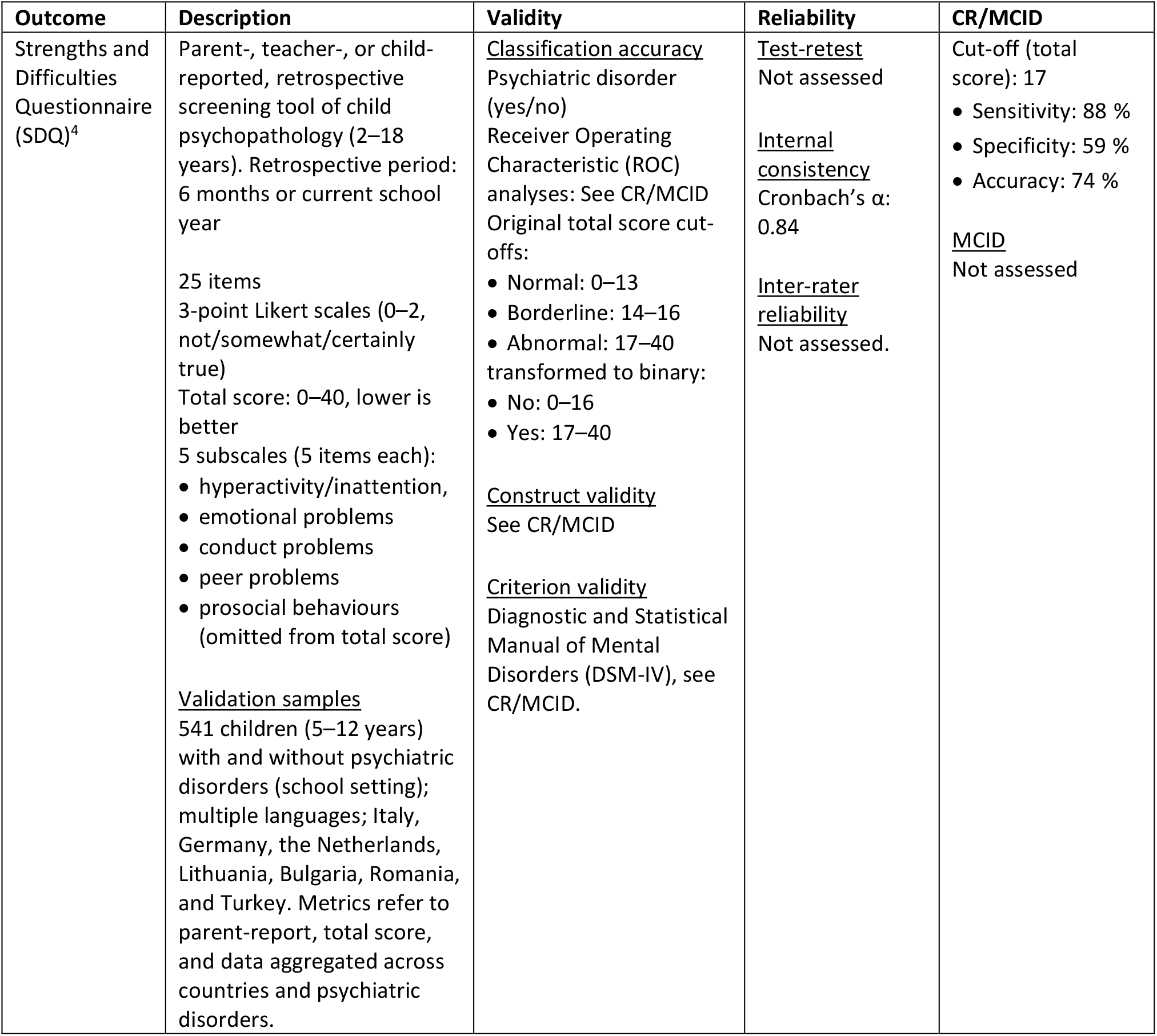
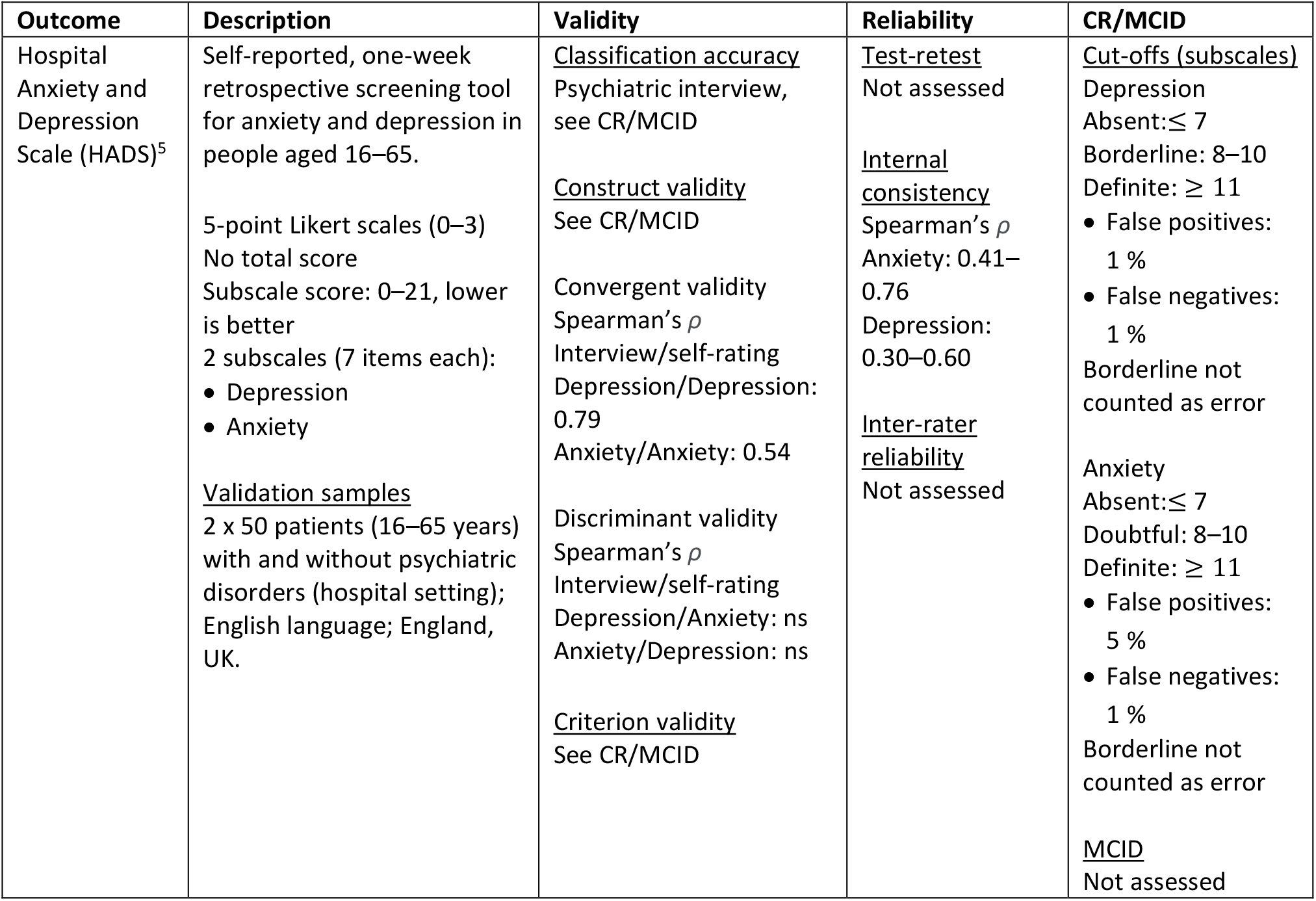
Psychometrics and clinical relevance/minimal clinically important difference (CR/MCID) for CASTLE Sleep-E outcomes. Metrics refer to the referenced publication, total scores and clinical samples unless otherwise stated.

| Outcome | Description | Validity | Reliability | CR/MCID |
| --- | --- | --- | --- | --- |
| Children's Sleep Habits Questionnaire (CSHQ)<sup>1</sup> | Parent-reported, one-week retrospective sleep screening tool for children (4–10 years)<br>35 items (2 duplicated across subscales)<br>3-point Likert scales (rarely, sometimes, usually)<br>Total score (33 items): 33–99, lower is better<br>8 subscales:<br>• Bedtime Resistance (6 items)<br>• Sleep Onset Delay (1 item)<br>• Sleep Duration (3 items)<br>• Sleep Anxiety (4 items)<br>• Night Wakings (3 items)<br>• Parasomnias (7 items)<br>• Sleep-Disordered Breathing (3 items)<br>• Daytime Sleepiness (8 items)<br><u>Validation samples</u><br>Parents of 469 school children (community setting) and 154 children diagnosed with sleep disorder (hospital setting); English language; England, UK.<br>Test-retest: 60 parents from control sample | <u>Classification accuracy</u><br>Sleep disorder (yes/no)<br>Receiver Operating Characteristic (ROC) analyses: See MCID<br><u>Construct validity</u><br>See MCID<br><u>Criterion validity</u><br>Not assessed | <u>Test-retest</u><br>2-week delay<br>Pearson's $r$: 0.62–0.79<br><u>Internal consistency</u><br>Cronbach's $\alpha$<br>Control sample: 0.68<br>Clinical sample: 0.78<br><u>Inter-rater reliability</u><br>Not originally assessed. | Cut-off (total score): 41<br>• Sensitivity: 80 %<br>• Specificity: 72 %<br>• Accuracy: 80 %<br><u>MCID</u><br>Not assessed. |
| Insomnia Severity Index (ISI), patient version<sup>2</sup> | Self-reported, one-month retrospective screening tool for insomnia in adults ($\geq 18$ years)<br>7 items<br>5-point Likert scales (0–4, no problem to severe problem)<br>Total score: 0–28, lower is better<br>• 0–7: Absence of insomnia<br>• 8–14: Subthreshold insomnia<br>• 15–21: Moderate insomnia<br>• 22–28: Severe insomnia<br>Dimensions:<br>• Severity of sleep onset<br>• Sleep maintenance<br>• Early morning awakening problems<br>• Sleep dissatisfaction<br>• Interference of sleep difficulties with daytime functioning<br>• Noticeability of sleep problems by others<br>• Distress caused by the sleep difficulties<br><u>Validation samples</u><br>959 adults with and without insomnia (community setting), 183 adults with insomnia and 62 controls (clinical setting); English language; Québec, Canada. | <u>Classification accuracy</u><br>Insomnia (yes/no)<br>ROC analyses, see MCID<br><u>Construct validity</u><br>See CR/MCID<br>Pearson's *r*<br>• Daily sleep diary: 0.54–0.59<br>• Activity level, Anxiety (state, trait), Depression, Fatigue (general, physical, mental), Motivation: 0.20–0.48<br><u>Criterion validity</u><br>Pearson's *r*<br>Polysomnography<br>• Sleep onset latency: ns<br>• Wake after sleep onset: ns<br>• Number of awakenings: ns<br>• Early morning awakening: ns<br>• Total wake time: ns<br>• Sleep efficiency: -0.16 | <u>Test-retest</u><br>Not assessed<br><u>Internal consistency</u><br>Cronbach's $\alpha$, Control sample: 0.71<br>Clinical sample: 0.73<br><u>Inter-rater reliability</u><br>Not assessed | <u>Control sample (self-diagnosis)</u><br>Cut-off (total score): 10<br>• Sensitivity: 86 %<br>• Specificity: 88 %<br>• Accuracy: 87 %<br><u>Clinical sample</u><br>Cut-off (total score): 11<br>• Sensitivity: 97 %<br>• Specificity: 100%<br>Accuracy: 98 %<br><u>MCID</u><br>Change required for improvement<br>Blinded assessor, *M*, [*CI*<sub>95</sub>]:<br>• Slight: 4.65 [2.61–6.69]<br>• Moderate: 8.36 [7.20–9.53]<br>• Marked: 9.89 [8.74–11.04]<br>ROC analyses:<br>• Slight: not reported<br>• Moderate: $\geq 7$<br>○ Sensitivity: 60 %<br>○ Specificity: 70 %<br>○ Accuracy: not reported<br>• Marked: $\geq 8$<br>○ Sensitivity: 64 %<br>○ Specificity: 80 %<br>○ Accuracy: not reported |
| Knowledge About Sleep in Childhood (KASC) | Self-reported scale assessing parental efficacy in managing child sleep and knowledge about child sleep<br>10 items | Not evaluated (custom-scale devised for CASTLE Sleep-E) | Not evaluated (custom-scale devised for CASTLE Sleep-E) | Not evaluated (custom-scale devised for CASTLE Sleep-E) |
| Health-Related<br>Quality Of Life Measure for Children with Epilepsy (CHEQOL)<sup>3</sup> | Quality of life assessment tool for children or parents with epilepsy (no specified time-period); child reported if $\geq 8$ years, parent proxy-report if child 5 to $< 8$ years<br>25 items<br>4-point Likert scales (0–4, opposites: true/sort of true)<br>Total score: 25–100, higher is better<br>5 subscales (5 items each):<br>• Interpersonal/social consequences<br>• Future worries<br>• Present worries<br>• Intrapersonal/emotional<br>• Epilepsy secrecy<br><u>Validation samples</u><br>381 children (6–15 years) with epilepsy and their parents (clinical setting); English language; Ontario, Canada. Test-retest: Additional 89, then 31 children; additional 48 parents.<br>Metrics refer to self-report for children 8–15 years and parent proxy report for children 5 to $< 8$ years and were assessed for sub-scales, not total score. | <u>Classification accuracy</u><br>Not assessed.<br><u>Construct validity (child)</u><br>Pearson's $r$<br>• Health care utilisation: 0.13–0.31<br>• Drug Adverse Events: 0.18–0.25<br>• Number of friends: 0.18<br>• N° of extracurricular activities: 0.13<br>One-way ANOVA ($p \leq .05$)<br>• Seizure severity: All 5 subscales<br>• Anti-epileptic drug use: 4 subscales<br>$t$-tests ($p \leq .05$)<br>• Help at school: All 5 subscales<br>Results for parent-proxy similar<br><u>Criterion validity</u><br>Not assessed | <u>Test-retest</u><br>10– 14 days delay<br>Intraclass correlation coefficient<br>Child: 0.59–0.69<br>Parent: 0.60–0.81<br><u>Internal consistency</u><br>Cronbach's $\alpha$, subscales<br>Child: 0.63–0.84<br>Parent: 0.64–0.86<br><u>Inter-rater reliability</u><br>Pearson's $r$<br>• Child/mother: 0.24–0.56<br>• Child/father: 0.18–0.54<br>• Mother/father: 0.40–0.71 | Not assessed. |
| Strengths and Difficulties Questionnaire (SDQ)<sup>4</sup> | Parent-, teacher-, or child-reported, retrospective screening tool of child psychopathology (2–18 years). Retrospective period: 6 months or current school year<br>25 items<br>3-point Likert scales (0–2, not/somewhat/certainly true)<br>Total score: 0–40, lower is better<br>5 subscales (5 items each):<br>• hyperactivity/inattention,<br>• emotional problems<br>• conduct problems<br>• peer problems<br>• prosocial behaviours (omitted from total score)<br><u>Validation samples</u><br>541 children (5–12 years) with and without psychiatric disorders (school setting); multiple languages; Italy, Germany, the Netherlands, Lithuania, Bulgaria, Romania, and Turkey. Metrics refer to parent-report, total score, and data aggregated across countries and psychiatric disorders. | <u>Classification accuracy</u><br>Psychiatric disorder (yes/no)<br>Receiver Operating Characteristic (ROC) analyses: See CR/MCID<br>Original total score cut-offs:<br>• Normal: 0–13<br>• Borderline: 14–16<br>• Abnormal: 17–40<br>transformed to binary:<br>• No: 0–16<br>• Yes: 17–40<br><u>Construct validity</u><br>See CR/MCID<br><u>Criterion validity</u><br>Diagnostic and Statistical Manual of Mental Disorders (DSM-IV), see CR/MCID. | <u>Test-retest</u><br>Not assessed<br><u>Internal consistency</u><br>Cronbach's $\alpha$: 0.84<br><u>Inter-rater reliability</u><br>Not assessed. | Cut-off (total score): 17<br>• Sensitivity: 88 %<br>• Specificity: 59 %<br>• Accuracy: 74 %<br><u>MCID</u><br>Not assessed |
| Hospital Anxiety and Depression Scale (HADS)<sup>5</sup> | Self-reported, one-week retrospective screening tool for anxiety and depression in people aged 16–65.<br>5-point Likert scales (0–3)<br>No total score<br>Subscale score: 0–21, lower is better<br>2 subscales (7 items each):<br>• Depression<br>• Anxiety<br><u>Validation samples</u><br>2 x 50 patients (16–65 years) with and without psychiatric disorders (hospital setting); English language; England, UK. | <u>Classification accuracy</u><br>Psychiatric interview, see CR/MCID<br><u>Construct validity</u><br>See CR/MCID<br><u>Convergent validity</u><br>Spearman's $\rho$<br>Interview/self-rating<br>Depression/Depression: 0.79<br>Anxiety/Anxiety: 0.54<br><u>Discriminant validity</u><br>Spearman's $\rho$<br>Interview/self-rating<br>Depression/Anxiety: ns<br>Anxiety/Depression: ns<br><u>Criterion validity</u><br>See CR/MCID | <u>Test-retest</u><br>Not assessed<br><u>Internal consistency</u><br>Spearman's $\rho$<br>Anxiety: 0.41–0.76<br>Depression: 0.30–0.60<br><u>Inter-rater reliability</u><br>Not assessed | <u>Cut-offs (subscales)</u><br>Depression<br>Absent: $\leq 7$<br>Borderline: 8–10<br>Definite: $\geq 11$<br>• False positives: 1 %<br>• False negatives: 1 %<br>Borderline not counted as error<br>Anxiety<br>Absent: $\leq 7$<br>Doubtful: 8–10<br>Definite: $\geq 11$<br>• False positives: 5 %<br>• False negatives: 1 %<br>Borderline not counted as error<br><u>MCID</u><br>Not assessed |

**Supplemental Table 3.** Composition, roles and responsibilities of the Trial Management Group, Programme Steering Committee, and Independent Data and Safety Monitoring Committee for CASTLE Sleep-E.

| Role | Name (Initials) | Affiliation |
| --- | --- | --- |
| **<u>Trial management Group (TMG)</u>** |  |  |
| **Responsibilities:** Day-to-day running and management of the trial. |  |  |
| **Meeting frequency:** Bi-weekly to three-monthly, depending on trial stage. |  |  |
| 1. King's College Hospital Sponsor Representative | Rahman Ahmed | King's College Hospital NHS Foundation Trust |
| 2. Chief Investigator | Deb K. Pal | King's College London, UK |
| 3. Co-Chief Investigator | Paul Gringras | Evelina London Children's Hospital, UK |
| 4. Co-Investigator<br>Public and Patient Involvement Lead | Lucy Bray | Edge Hill University, UK |
| 5. Co-Investigator<br>Qualitative Research Lead<br>Public and Patient Involvement Co-Lead | Bernie Carter | Edge Hill University, UK |
| 6. Co-Investigator<br>Health Economics Lead | Dyfrig Hughes | Bangor University, UK |
| 7. Co-Investigator<br>Patient Reported Outcome Lead<br>Public and Patient Involvement Co-Lead | Christopher Morris | University of Exeter, UK |
| 8. Co-Investigator<br>Lead Statistician | Catrin Tudur Smith | University of Liverpool, UK |
| 9. Co-Investigator<br>Intervention Development Lead | Luci Wiggs | Oxford Brookes University, UK |
| 10. Supervising Trials Manager | Catherine Spowart | University of Liverpool, UK |
| 11. Trial Manager | Lucy Stibbs-Eaton | University of Liverpool, UK |
| 12. Trial Statistician | Victoria Watson | University of Liverpool, UK |
| 13. CASTLE Programme Manager | Amber Collingwood | King's College London, UK |
| 14. Researcher | Georgia Cook | Oxford Brookes University, UK |
| 15. Researcher | Kristina C. Dietz | King's College London, UK |
| 16. Health economist | Will A. S. Hardy | Bangor University, UK |
| 17. Researcher | Holly Saron | Edge Hill University, UK |
| **<u>Trial Steering Committee (TSC)</u>** |  |  |
| **Responsibilities:** Overall trial supervision and advice, ultimate decision for the continuation of the trial. |  |  |
| **Meeting frequency:** At least annually. |  |  |
| 1. Chair | Jeremy Parr | Newcastle University, UK |
| 2. Medical statistician | Martyn Lewis | Keele University, UK |
| 3. Paediatrician | Desaline Joseph | Evelina London Children's Hospital, UK |
| 4. Public and Patient Involvement Representative | Jo Conduit-Smith | CASTLE Advisory Panel |
| **Independent Data and Safety Monitoring Committee (IDSMC)**<br>**Responsibilities:** Interim monitoring of safety and effectiveness, trial conduct and external data.<br>Recommendation to TSC about trial continuation.<br>**Meeting frequency:** At least annually |  |  |
| 1. Chair | Helen Cross | University College London, UK |
| 2. Paediatrician | Alberto Verroti | University of L'aquila, Italy |
| 3. Medical statistician | Anthony Johnson | University College London, UK |

**Supplemental Table 4.**
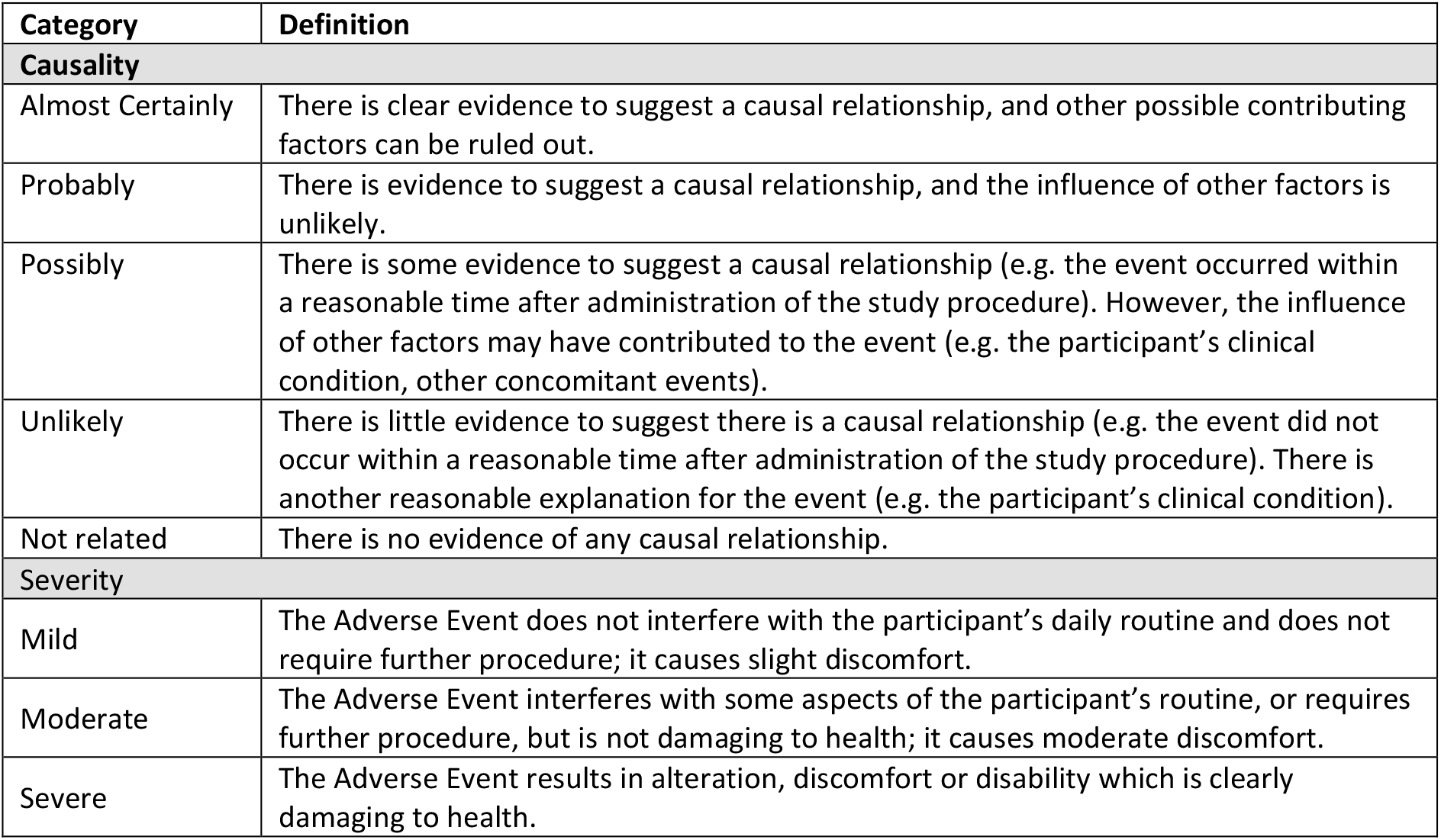
Categories used to define the causality and severity of Adverse Events in CASTLE Sleep-E.

**Supplemental Figure 1.**
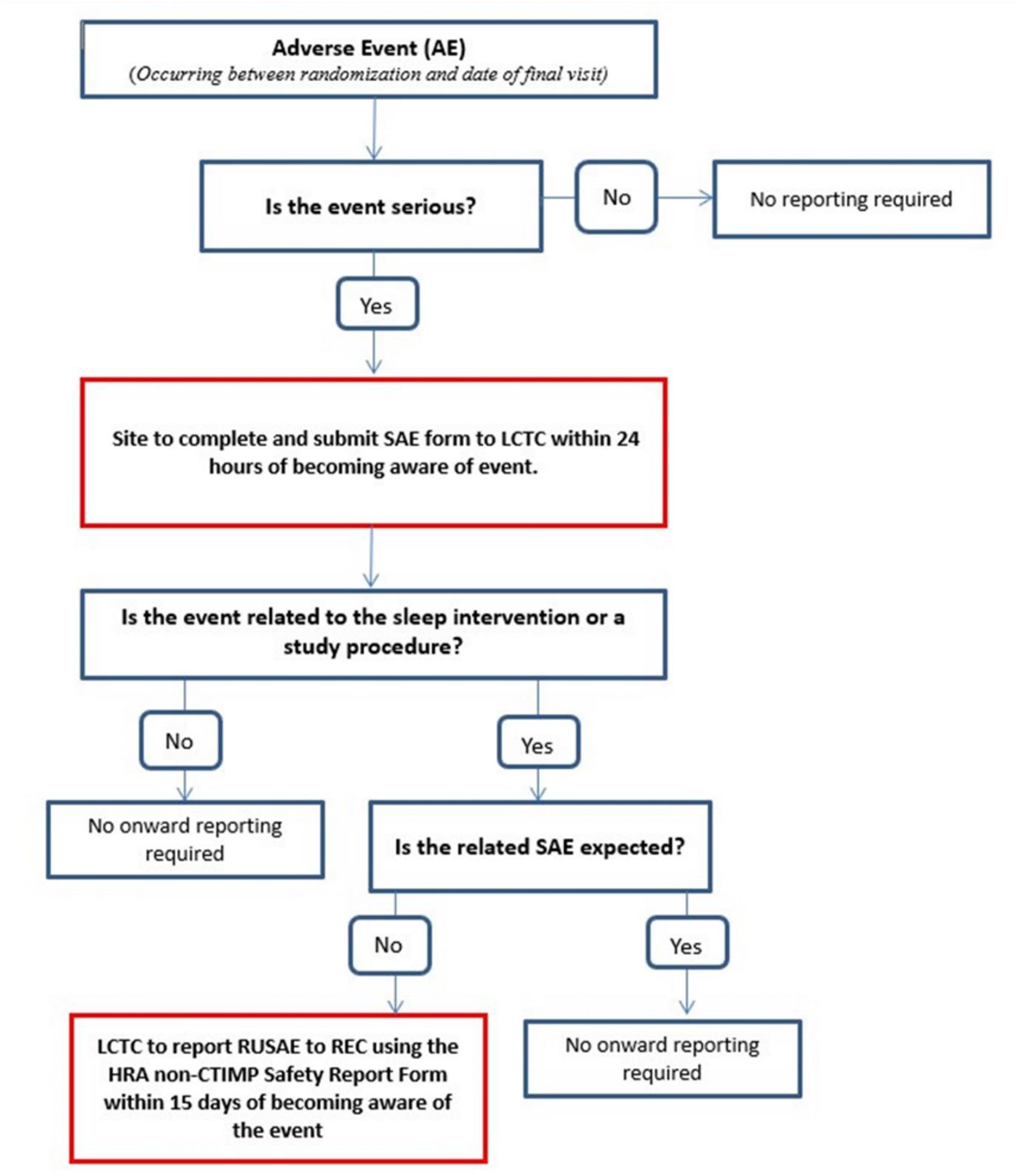
Flowchart showing reporting requirements of Adverse Events for the CASTLE Sleep-E trial. Acronyms: Serious Adverse Event (SAE), Liverpool Clinical Trial Centre (LCTC), Related Unexpected Serious Adverse Event (RUSAE), Health Research Authority (HRA), non-Clinical Trial of Investigational Medicinal Products (non-CTIMP).

## Parent/Guardian Information Sheet for CASTLE Sleep-E

- You have been given this information sheet as your child might be eligible to take part in this research study. Please take time to read the following information carefully.
- Part 1 tells you the purpose of the study and what taking part means for you and your child.
- Part 2 gives you more detailed information about who is running the study and what happens to your data.
- CASTLE Sleep-E is a study for children with Rolandic epilepsy (RE) who are experiencing sleep problems.
- the study we will compare an online sleep intervention (known as COSI) against standard care
- If your child is aged between 5 years and 12 years then you and your child might be eligible to join.
- We will recruit 110 children and their families across the UK.
- All parents and children in the study will be asked to wear an actigraph (a watch-like device that records your sleep) for 2 weeks at 2 different times during the study.
- All families will be asked if they are happy to be interviewed by a researcher, however not all families who express an interest in these interviews will be contacted. If there is anything that is not clear, or you would like more information, please ask a member of the clinical team.
- If you wish, you can discuss the study with friends, relatives and/or get independent advice via your local Patient Advice and Liaison Service (PALS) or equivalent.

### How to contact the study team

If you have any questions about this study, please talk to your research team:

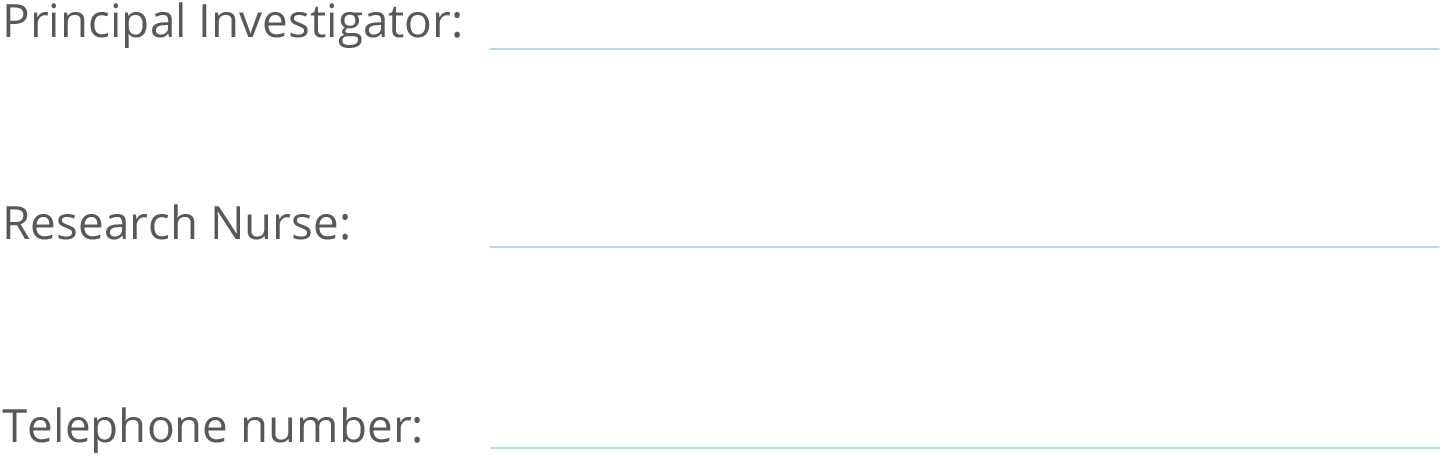

## Part 1 Purpose of the study and what will happen if your child takes part

### 1.1 Why are we doing the CASTLE Sleep-E study?

Epilepsy is a common condition among children in the UK. Families have identified sleep problems in their children with epilepsy and also amongst parents as a major issue that doesn’t get enough attention. Rolandic epilepsy is the most common type of epilepsy in childhood, also sometimes known as ‘benign Rolandic epilepsy’ ‘or ‘childhood epilepsy with centrotemporal spikes’. Children with Rolandic epilepsy often have seizures at night and their seizures can be triggered by poor sleep. Sleep problems can be present while they are being followed up by their paediatrician for seizures and even persist after the seizures have gone away. Sometimes their learning, behaviour, self-esteem and mood are affected too.

Sleep problems can be managed through practice. There are guidelines to help children in general with their sleep, but there is nothing available that specifically helps children with epilepsy and their parents address sleep problems and improve their sleep quality.

Therefore the CASTLE Sleep-E study aims to find out whether giving families access to a newly developed online sleep intervention (known as the CASTLE Online Sleep Intervention or “COSI” for short) will help improve their quality of sleep. We will compare the child and parent’s sleep quality at the start and after three months. In order to make a fair and balanced comparison, half the families will receive COSI and the other half will receive standard care from their paediatrician. With these equally divided groups we’ll be able to evaluate if COSI works or not, as well as review which treatment makes the best use of NHS resources and how these treatments affect your and your child’s quality of life.

The information given in COSI is personalised depending on the answers provided by the family and has been designed specifically for parents of children with epilepsy. Not all families who enter CASTLE Sleep-E will receive access to COSI. Some may be treated using standard care, which will mean they will not receive access to the system.

Families who are happy to take part in CASTLE Sleep-E will be followed up for 6 months. If you are happy for your child to be part of the study, you and your child will be asked to wear an actigraph (sleep monitor) for 2 weeks and complete some questionnaires and assessments before your child is allocated to their study group. You and your child will also be asked to wear the sleep monitor and complete an assessment again 3 months after your child is allocated their study group. We will also ask you and your child to complete some questionnaires electronically 3 and 6 months after your child has received their study group. Your local research team will collect information about you and your child at the same timepoints. We collect information at different times during the study so we can compare how your child was before and after they are allocated their study group.

We will collect your and your child’s questionnaire answers on a secure website, the link to which will be emailed to you. You will also be given an iPad to use during your time on the study. The iPad will be sent directly to your home and you will only be able to access Apps required for the study. It will be used to complete your child’s “SleepSuite” assessment and may also be used to complete the study questionnaires and access COSI (if allocated to that study group).

Sleepsuite is an iPad adventure game that children complete before bedtime, and then the next morning. It’s been designed to capture changes in things like word memory and reaction time that we know improve overnight after a night’s sleep.

There is also an opportunity to take part in two interviews during the study, these will be carried out 3 months and 6 months after your child is allocated their study group. If you do not wish to be interviewed, this will not impact your child taking part in the main study.

When the CASTLE Sleep-E study ends, access to the online sleep intervention used as part of the study will be closed but all families will be given the option to receive the written content of COSI to keep. This will be sent to you in an electronic format via email (if you wish to receive it).

The results from this study will be used to help us improve treatments for children with epilepsy.

### 1.2 Why has my child been invited?

Your child has been invited to take part in CASTLE Sleep-E because they are between the ages of 5 and 12 with diagnosed Rolandic epilepsy and are having problems with their sleep.

### 1.3 Does my child have to take part?

No, taking part is voluntary. It is up to you to decide whether or not your child should take part.

If you and your child decide not to take part, then your child will still receive the usual treatment their hospital offers. Their doctor can provide you with more information on this. If you decide that you and your child will take part, you can change your mind at any time later without giving a reason.

The decision you make on whether your child will take part or not doesn’t affect the standard of medical care they receive now or in the future

### 1.4 What organisations are involved in this study?

The CASTLE Sleep-E trial involves a number of different activities which are carried out by different organisations across the UK. The below table will provide a bit more information about these organisations and the study activities they are responsible for.

| Name of organisations | CASTLE Sleep-E Activities |
| --- | --- |
| Liverpool Clinical Trials Centre (also known as "LCTC") | The Liverpool Clinical Trials Centre is responsible for the day-to-day management of the CASTLE Sleep-E study. They ensure the trial is carried out in line with strict standards.<br>LCTC is responsible for collecting and reviewing clinical and questionnaire data. At the end of the study, the team at LCTC will review whether COSI (sleep intervention) has been effective. |
| Bangor University | Bangor University is responsible for conducting the health economics analysis which will assess the cost-effectiveness of the COSI compared to standard care. |
| Edge Hill University | The team at Edge Hill University is responsible for carrying out interviews with families (if they consent to this activity) to find out about family's experiences with Rolandic epilepsy and sleep. |
| Oxford Brookes University | Oxford Brookes University manage the actigraphy (sleep monitor) activity within CASTLE Sleep-E. The Sleep Team will be in contact with participants at a number of different times during the trial to arrange delivery and collection of study devices, such as actigraphs and iPads. |

Further information about the different study activities mentioned above can be found in section 1.5 of this information sheet.

### 1.5 What does taking part involve for my child and me?

If you agree for your child to take part, you will first be asked to sign a consent form electronically. Signing this consent form will be completed following a discussion with your local research team. The research team will provide you with a paper or electronic copy of the completed consent form after your visit or telephone/video call.

Your local study team will then check and confirm that this study is suitable for your child, and you and your child will be asked to follow the study plan. Your child will be enrolled in the study for 6 months. During your time in the study, you and your child will be followed up by your paediatrician and research/ specialist nurse. You and your child will also be asked to do the following:

- Complete some electronic questionnaires
- Wear a wrist actigraph (sleep monitor) for two weeks at the start and again part way through the study (at 3 months)
- Your child will complete a special iPad game called SleepSuite at the start of the study and 3 months after their treatment allocation.

We will email a link to the electronic questionnaires when they are due to be completed, which you can access using an electronic device (for example, a smartphone, tablet, or computer). The “SleepSuite” task must be completed on the study iPad, not on any other device. The iPad and wrist actigraph will be delivered directly to your home by courier.

Research nurses will collect information about your child’s health and well-being at your clinic appointments with your paediatrician (which will be held either face to face in clinic or via telephone/ video call depending on your local hospital and the coronavirus situation).

If you are happy to be interviewed as part of the study, you and your child might be selected to take part in two extra telephone or video-calls with a researcher.

Further information about the study visits can be found below.

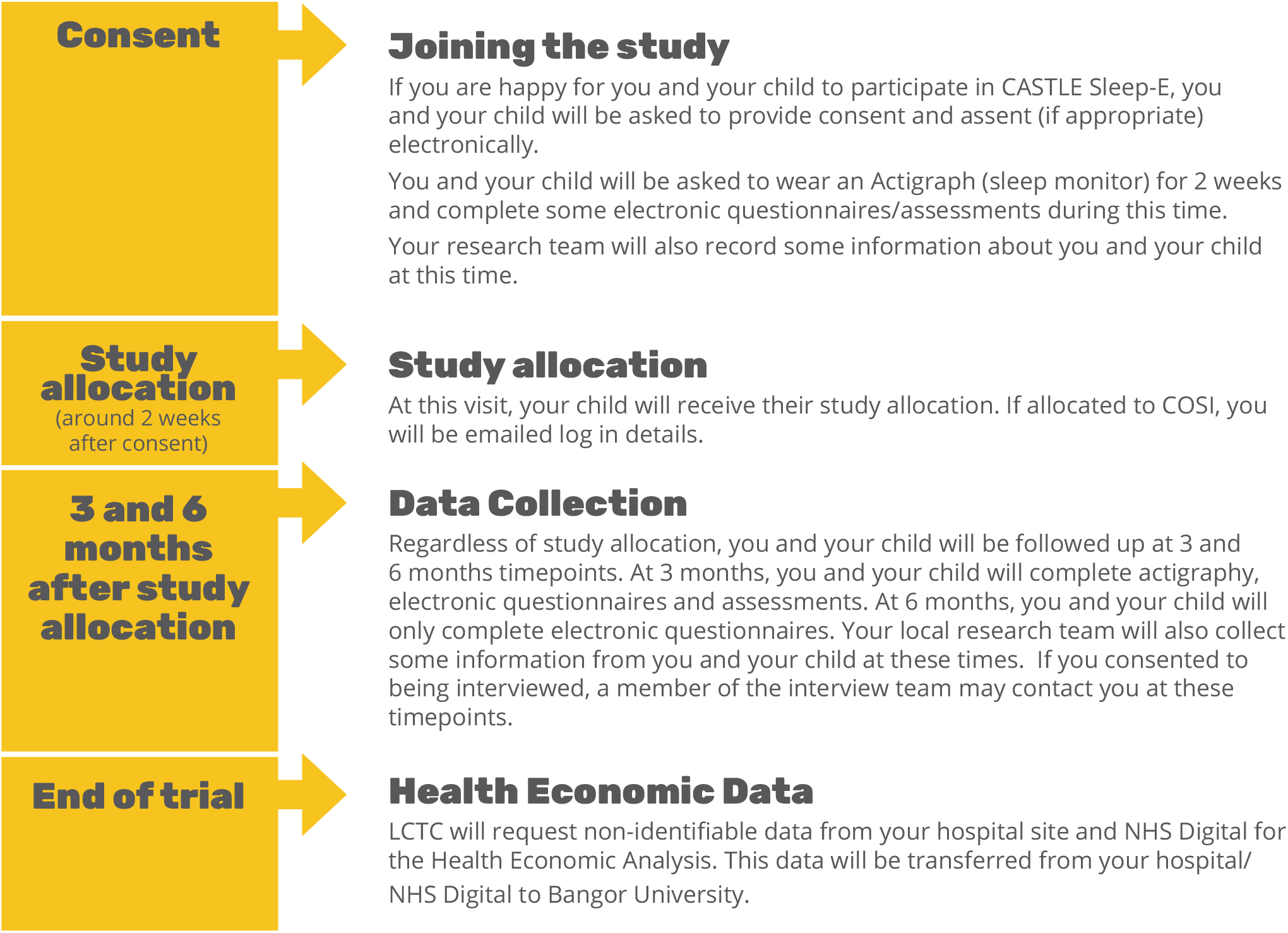

#### Consent Visit (Visit 1)

This visit will be with a member of your hospital research team and may be face-to-face in clinic or via a telephone/video call. It will take around 1 hour. It is important to know that your child’s study group will not be decided at this visit.

Your research team will ask you some questions at this visit about you and your child. They will also look at your child’s EEG results. The questions asked and EEG results will help confirm if this study is right for you and your child.

You will be able to ask any questions that you may have about the study. If you have had all of your questions answered and you are happy for your child to take part, then you will be asked to sign a consent form electronically.

You will receive an email with a secure link to the consent form which you can sign in clinic or at home, depending where your visit is held. You will be asked to provide your contact details as part of the consent form, which includes your name, mobile telephone number, email address and home address (including postcode).

If your child is 7 years or older, we will also ask if he/she is happy to take part in the study and to sign an assent form electronically by writing their name, if they can.

The researchers at site will provide you with a copy of the completed consent form along with the child’s completed assent form (if one has been completed) after your visit or telephone/video call.

As part of the consent form, you will be asked to consider how you’d like your and your child’s information to be used. Bangor University, who are undertaking the Health Economic analysis, will request consent to use information collected during the study as well as routinely collected NHS data for patients taking part in CASTLE Sleep-E. The data collected during the study will be transferred by LCTC to Bangor University. The data collected at the end of the study will be transferred directly to Bangor University by NHS Digital and the NHS hospital that your child attends. In both instances, the data will be coded prior to transfer so that it is not possible to identify you or your child.

At this visit you will also be asked if you are happy to be interviewed by the qualitative research team based in Edge Hill University. This is optional and if you do not wish to be interviewed, you can still join the main study. If you are happy to be interviewed, you will be asked to give your consent and for your child to assent (if appropriate). You may be contacted 3 months and 6 months after your child has received their study group allocation. Only around half of all families taking part in the main part of the CASTLE Sleep-E study will be selected for interview but we would be grateful if you could take part.

During this first visit your doctor or research nurse will give you information relating to your sleep monitor, study iPad and questionnaires. There is more information about wearing a sleep monitor, your study iPad and the questionnaires you need to complete below.

#### Wearing a sleep monitor (actigraph)

Children who take part in CASTLE Sleep-E and one of their parents will be asked to wear a sleep monitor for 14 days (2 weeks) at 2 different times in the study. After signing your consent form, you and your child will be asked to wear a sleep monitor for 14 days once you receive it. After that period your child will be allocated to one of the two study groups.

The sleep team for CASTLE Sleep-E (who are based at Oxford Brookes University) will receive your contact details after you consent to the study and will give you a call to arrange a suitable delivery time for the sleep monitor and study iPad.

You will be provided with instructions for the sleep monitor and how to return it (at no cost to you), as well as the sleep team’s contact details in case you have any queries. You will also be asked to complete a sleep diary for you and your child whilst you are wearing the monitors. This is to help us interpret the information recorded by the monitors.

#### The study iPad

The sleep team will send you a study iPad at the same time as the sleep monitors. There is no cost to you to use the iPad, but we ask that you keep it safe as you will need to complete your child’s “SleepSuite” assessments on it twice during the study. The iPad will be specially set up to limit access to other apps. You will be asked to complete your child’s assessments on this iPad at two different times during the study. You will be provided with instructions on how to use the iPad.

At the end of your child’s participation in the study, you must return the study iPad, charger and cable to your local research team at your next visit. If you are unable to leave your study iPad with your local research team, a member of the CASTLE Sleep-E team will contact you to arrange an alternative solution.

#### Questionnaires

Before your child is allocated to their study group, you and your child will be asked to complete some questionnaires. These questionnaires are completed electronically and will be sent to you via email in a secure weblink. The web link can be accessed on an electronic device with access to your emails, such as a computer, smartphone or tablet. You may also use the study iPad to complete these questionnaires. These questionnaires should be completed in the same 2 week period that you and your child are wearing the sleep monitors. You will receive a reminder about completing the questionnaires one week after you receive your actigraph. If you have any difficulties with the questionnaires, please contact a member of your local research team.

All the questionnaires included in CASTLE Sleep-E are being used as research tools. They are only analysed at the end of the study, and the data are analysed as ‘groups’ rather than as individuals. The CASTLE Trial Management Group are therefore unable to comment clinically or otherwise feedback on individual results – either child or carer. If you have concerns about any issues raised in these questionnaires, please discuss them with your local research team or GP.

#### Randomisation Visit (Visit 2)

This appointment will take place once you and your child have worn your sleep monitors for 2 weeks. The appointment will take place a maximum of 4 weeks after your first visit and will be held via telephone/video call, unless your research team have specifically requested a face-to-face visit. This appointment will take around 30 minutes. During this appointment, a member of your research team will confirm you are still happy to take part in the study and check your contact details are correct.

If you are happy to take part, you will be asked some questions about your child’s general health, their seizures, any medication they may be taking and any school absences they may have had due to their epilepsy. Once these questions have been answered, your child will be allocated into one of the two study groups (sleep intervention or standard care) using a computer programme. Your doctor or research nurse will tell you which group your child has been allocated to.

#### Accessing the sleep intervention

If your child is allocated to the group which receives sleep intervention, you will receive an email with a link to a website called “COSI” (Castle Online Sleep Intervention). The email will also contain the login details (username and password) to access your account in the COSI website. You will be able to access the system from any electronic device from which you access your emails (such as a computer, tablet or smartphone). As with the study questionnaires, you may wish to use the study iPad to access COSI.

Once you are logged in, you will have access to information, including a series of videos and explanations that will give you sleeping strategies/tips to apply to your child’s sleep to try and improve their sleep, as well as yours.

If you don’t access COSI within the first couple of days after the account has been created, you will receive an automated text reminder, which will be followed up by an email reminder if the system is still not accessed within a further few days. If the COSI system is not accessed within 6 days of the account being created, you will receive a quick telephone call from a member of the CASTLE Sleep-E team.

The CASTLE Sleep-E team will contact you again by telephone 6 weeks after the account is created to discuss COSI and how you are finding using the system. The team will be unable to offer any specific clinical advice during this phone call.

3 months after your account is created, you will be contacted by email and asked to complete an online evaluation questionnaire about COSI. If you don’t complete this questionnaire within a few days, you will receive an automated text reminder which will be followed by a telephone call from the sleep team if the evaluation questionnaire has still not been completed.

#### Follow-up visits at 3 months and 6 months (Visits 3 and 4)

You and your child will be followed up by your paediatrician and research nurse 3 and 6 months after your child is allocated to their study group. These follow up visits may take place face-to-face in clinic or via a telephone/video call. Each follow up visit will take around 30 minutes.

During these visits, your local research team will ask you if you are still happy to remain in the study and if so, will ask questions around your child’s seizures, any medication they may be taking and their general well-being, including any school absences they have had. A member of the research team will also check your contact details are correct.

Around the time of your visits, you will also be sent an email from the CASTLE Sleep-E team asking you and your child to complete some questionnaires and assessments on the study iPad (the iPad “SleepSuite” assessment will only be completed at 3 months). You and your child will also be asked to wear your sleep monitors for 2 weeks at 3 months.

If you gave consent at Visit 1 for an interview, you may be contacted a few weeks after your follow up visits by the interview team at Edge Hill University. Further information about the interviews can be found below.

#### Interviews

If you are selected, then you will be invited to take part in two interviews. The first one will be a few weeks after Visit 3 and the second one a few weeks after Visit 4. We will arrange the interviews at a time to suit you (and your child).

The interviews will be done remotely using a secure video-calling system. Although we will only record the audio part of the interview, we hope you will be happy to use the video option when we are talking. Each interview is likely to last between 30-60 minutes but this can be tailored to suit how much time you have available and how much you want to share with us. We will send you a list of the sorts of questions we expect to ask you about a week before the interview. We hope this will help you feel confident in the interview.

If your child is aged 7 years or older, they will also be able to take part in the interview study. The interviews will be done by video-call. We plan to send your child an activity booklet about a week before we talk to them. We will send the activity booklet by email and/or post, as you prefer. The booklet or sheet will help them think about what they would like to tell us. The booklet/sheet will give your child a chance to share their ideas in different ways (for example, using pictures, writing down their ideas, choosing emojis to represent their feelings). If your child would prefer just to talk to us, they don’t need to fill in the booklet/sheet.

We hope you will share a copy of the completed booklet/sheet with us before the interview by email (or other suitable means) or share it at the interview by showing it to us.

The sorts of things we are likely to ask you and/or your child questions about include:

- Your experiences of living with Rolandic epilepsy (including what you think about your child’s seizures, treatment and how your ideas might change over time);
- Your experiences of your child’s sleep issues and any impact that this has on your family and how you manage them;
- experiences of using COSI, wearing the sleep monitors and what you liked and what we could improve; and
- How you make decisions about your child’s epilepsy, their sleep and using COSI.

We will audio-record your interview. The audio recordings of the interviews will be sent to be typed up by a third-party transcription company. This company will remove any information from the transcriptions which may identify you or your child. This company have signed a confidentiality agreement with Edge Hill University and will not use your recording for anything other than the purposes of the study. The recording of your interview will be deleted one week after they have been typed up and the researcher has checked that we have an accurate record of the interview. If you would like, we can send you a summary of the key things we talked about in your interview. We will use the typed-up version of the interview as the basis for our analysis.

With your and your child’s consent, we will keep a copy of their activity booklet. We would also like your and your child’s consent to use their drawings and responses to the activities (including direct quotes) in our reports and presentations. We will remove anything that will identify you or your child.

#### Postcards

With your permission, we would like to send your child a postcard at three different times during the study. Two of the postcards will provide an update on the study, as well as a fun activity for your child. The final postcard will be sent with an accompanying certificate to thank them for taking part.

### 1.6 What exactly is COSI?

The CASTLE Online Sleep Intervention is known as “COSI” for short. The system is accessed using a web link, which will be emailed to those families allocated to the sleep intervention group.

The information in COSI gives parents advice about strategies that are helpful for improving the sleep of children, specifically designed to meet the needs of parents of children with epilepsy.

COSI includes information about many different types of sleep problems and gives ideas for things you can do to try and improve your child’s sleep and how to deal with common issues along the way, including some of the special issues faced by children with epilepsy and their families.

COSI is interactive; it will ask for some information about your child and their sleep so that COSI knows which type of information is likely to be of most use to you. Of course, you can look at any section in COSI but, to save you time, you can also look just at the sections which are highlighted as being relevant for you and your child’s sleep problems.

### 1.7 How will I know which study group my child is going to be in?

In research studies we often split patients up into groups to look at how different treatments or interventions work. In the CASTLE Sleep-E study patients will be split into two study groups. Neither the families or the researchers get to choose who goes in which study group and this is to create fair and balanced groups of children and parents in each group.

- One group will receive access to the online sleep intervention (known as the CASTLE Online Sleep Intervention or “COSI” for short)
- The other group will receive standard care from their paediatric team (no access to COSI)

It is really important that each group in the CASTLE Sleep-E study has a similar mix of patients in it so we know that if one group of patients does better than the other it is very likely to be because of the intervention and not because there are differences in the types of patients in each group.

We use a computer programme that puts patients into groups ‘at random’ – you might hear this described as ‘randomisation’ or ‘random allocation’, but they all mean the same thing. It means that neither you nor your child’s doctor choose who gets to be in which group so that the groups are fair and balanced.

In the CASTLE Sleep-E study your child is equally as likely to be in the group receiving online sleep intervention as they are in the group receiving standard care

### 1.8 What are the alternatives for treatment?

If you do not wish to take part in CASTLE Sleep-E, you will be treated as per standard care, which means your doctor will advise you on the course of treatment which is right for your child.

### 1.9 What are the benefits and risks of taking part?

Families who take part in CASTLE Sleep-E will receive standard NHS care for their time on the study. There are no additional risks to taking part in the study.

The main benefit anticipated from the sleep intervention (“COSI”) is improved sleep compared to standard care. It is possible that there may also be a reduction in seizures when following the sleep intervention compared to standard care. If, however, your child’s doctor decides that seizures are not being controlled well, they may discuss additional treatments (such as anti-seizure medicine) with you.

We hope that the results from the study will help doctors and patients in the future when making decisions about treatment.

### 1.10 What happens if I change my mind?

You and your child have the right to withdraw from the study at any time, without giving a reason. Your child can choose to withdraw independently of your wishes and in this instance, both you and your child will not take part in the study.

If at any point you or your child decide to stop taking part in the study, your child will still receive medical treatment and the follow up usually offered by their hospital.

If you do decide that you both should stop taking part, we will ask you if you and your child would like to:

- continue to complete follow up visits for the study or
- stop taking part with no more study visits.

Information on how we will handle your and your child’s information in the event of you both withdrawing is detailed in Part 2 of this Information Sheet.

### 1.11 What if new information becomes available?

Sometimes during the course of a research project, important new information becomes available about the treatment or intervention that is being studied. If this happens, the doctor will tell you about it and discuss with you whether you want your child to continue in the study. If you decide to withdraw your child your doctor will make arrangements for their care to continue. If you decide they should continue in the study you will be asked to sign an updated consent form.

On receiving new information, the doctor might consider it to be in your child’s best interests to withdraw them from the study. He/she will explain the reasons and arrange for their care to continue.

If the study is stopped for any other reason you will be told why and your child’s continuing care will be arranged.

### 1.12 What happens when the study stops?

When the CASTLE Sleep-E study ends, access to the online sleep intervention used as part of the study will be closed but all families will be given the option to receive the written content of COSI to keep. If COSI is found to be helpful, ultimately we plan to make an online version of COSI available to all families of children with epilepsy.

It is intended that the results of the study will be presented at conferences and published in medical journals so that we can explain to the medical community what our research results have shown. They may be used to apply to the necessary authorities to make the intervention widely available, if shown to be beneficial.

Confidentiality will be ensured at all times and neither you nor your child will be identified in any publication.

Any information derived directly or indirectly from this research, as well as any patents, diagnostic tests, drugs, or biological products developed directly or indirectly as a result of this research may be used for commercial purposes. Neither you nor your child have any right to this property or to any share of the profits that may be earned directly or indirectly as a result of this research. However, in signing this form your child does not give up any rights that they would otherwise have as a participant in research.

### 1.13 What if there is a problem?

Any complaint about the way you or your child have been dealt with during the study or any possible harm they might suffer will be addressed. Detailed information is given in Part 2 of this information sheet.

### 1.14 Will taking part in the study be kept confidential?

Yes. All the confidential information about your child’s participation in this study will be kept confidential. Detailed information on this is given in Part 2.

## Part 2 Detailed Information about the conduct of the study

### 2.1 Who is running the study?

King’s College Hospital NHS Foundation Trust and King’s College London are the Co-Sponsors of this study and are responsible for managing it. They are based in the United Kingdom. They have asked that the day to day running of the study is carried out by a team based at the Liverpool Clinical Trials Centre (LCTC, part of the University of Liverpool) with help from the Health Economics Researchers from Bangor University, Qualitative Researchers from Edge Hill University and the Sleep Team from Oxford Brookes University.

The study has been reviewed by the Health Research Authority and the East Midlands – Nottingham 1 Research Ethics Committee to make sure that the study is scientifically and ethically acceptable.

This study is publicly funded by the National Institute for Health Research (NIHR). Your child’s doctor will not receive any payment for including them in this study.

### 2.2 How will my and my child’s information be collected and handled?

King’s College London and King’s College Hospital NHS Foundation Trust are the joint Data Controllers for this study. Bangor University (along with King’s College London and King’s College Hospital) will be a joint data controller for the subset of NHS Digital data obtained at the end of the study. These organisations will need to use information from you, your child and his/her medical records for this research project.

This information will include:

- Your name
- Your child’s name/gender/initials/date of birth
- Your child’s NHS/CHI/H&C* number

*NHS number is only used in England and Wales. CHI number used in Scotland and H&C number in Northern Ireland.

- Your contact details (including address, postcode, mobile phone number and email address)

Members of the CASTLE Sleep-E research team from King’s College London, King’s College Hospital NHS Foundation Trust, LCTC and regulatory organisations will use this information to do the research or to check your child’s records to make sure that the research is being done properly. Members of the CASTLE Sleep-E research team may also look at your child’s medical and your and your child’s research records, and the consent form you sign, to check the accuracy of the research study. Bangor University will only view and use non-identifiable information.

People who do not need to know who you and your child are, will not be able to see your or your child’s name or contact details. Your and your child’s data will have a code number (study ID) instead.

#### Medical and study information

Your child’s NHS hospital will collect information from you, your child and his/her medical records for this research study in accordance with our instructions.

Your child’s NHS hospital will use the above listed contact information, along with your child’s study ID number, to contact you and your child about the research study. They will also use these data to make sure that relevant information about the study is recorded for your child’s care and to oversee the quality of the study.

The contact information collected on the consent form you complete will be securely stored by the Liverpool Clinical Trials Centre (LCTC), which is part of the University of Liverpool and coordinates the study. LCTC will then securely pass yours and your child’s contact details to Oxford Brookes University electronically. LCTC will also pass your mobile phone number onto a third-party text messaging company. This company will use your mobile phone number for study purposes only (namely the purpose of sending of automated text reminders) and will not share it more widely. LCTC may also securely pass your contact details to Edge Hill University, if you consent to being interviewed during Visit 1.

Your email address and mobile telephone number will also be collected to give you access to COSI (if allocated to this treatment arm) and send you email/ text and telephone reminders. Some of these text reminders will be automated and sent to you from a third party text messaging company (as mentioned in above). Your telephone number will be used to arrange the delivery of the sleep monitor and study iPad, which will be sent to your home address. To do this your name and mobile number will be stored on a passcode protected mobile phone which is used purely for the research project to help facilitate ease of communication between you and the research team.

The CASTLE Sleep-E team at Oxford Brookes University will transfer your name, postal address and telephone number to a reputable courier for the purposes of delivering and collecting your study iPad and Actigraph. The information provided to the courier will be used for study purposes only and not shared more widely.

The only people from the trial co-sponsors, LCTC, Oxford Brookes and Edge Hill University who will have access to information that identifies you/your child will be people who need to contact you/your child, audit the data collection process or share your information with NHS Digital for the purpose of health economic analysis.

The people who analyse the information at LCTC and Bangor University will not be able to identify you and your child and will not be able to find out your/his/her name, NHS/CHI/H&C number or contact details. Every effort will be made to ensure that any further information about you and your child that leaves her/his NHS hospital (captured on forms and databases used to collect data for the study) will have your/her/his name removed so that you/she/he cannot be recognised from it; this information will usually be removed by a member of the study team at your child’s NHS hospital, but may also be removed by LCTC upon receipt.

Pseudo-anonymised data (data that will not directly identify you) collected by researchers and from you and your child at the clinic visits and by questionnaires will be entered directly into the CASTLE Sleep-E study database, held by LCTC, for storage and analysis. Relevant data needed by other members of the research teams at Bangor University, Edge Hill and Oxford Brookes will also be held for analysis and storage.

With your consent, your contact details will be sent to the CASTLE Sleep-E team at Oxford Brookes University and Edge Hill University via a secure database (managed by LCTC). Oxford Brookes will use this information so they can send you the sleep monitors and iPads, as well as to contact you to discuss the sleep intervention (if allocated to this group). Edge Hill University will only be able to access and use this information if you agree to take part in the interview study.

If you and your child wear a sleep monitor, you will send the monitor back to the CASTLE Sleep-E team at Oxford Brookes University and then the data will be sent to the LCTC for analysis. The data will only be accessed by people working on the study or people working to ensure the study is being run correctly. If you log in to the COSI website, pseudo-anonymised data will be collected by LCTC on the usage of this website so we can see if it was useful.

When your child completes the iPad SleepSuite game, pseudo-anonymised data will be securely uploaded to a cloud (online backup) where only authorised CASTLE Sleep-E members from LCTC and King’s College London can access the information. At the end of the study, your child’s data will be removed from the cloud storage services and transferred to LCTC for analysis.

If you consent to being interviewed as part of the study, a recording of your/your child’s interview will be sent by Edge Hill University to a third-party transcription company who will type up your responses. Once a researcher from Edge Hill University confirm that the transcription is accurate, this recording will be deleted.

With your consent, we will send a letter to your child’s GP to let them know your child is taking part in the CASTLE Sleep-E study.

The routine Electroencephalography (EEG) reports (test that measures the electrical activity of the brain) will be sent for independent review by doctors at King’s College London working on the CASTLE Sleep-E study at the end of the study to assess the rolandic epilepsy diagnosis made by your local clinician. The reports will not have yours or your child’s personal details on them. As identifiable information will not be present on the EEG report, it will not be possible to feed back any findings to you.

At the end of the study, the Health Economics team working at Bangor University (courtesy of LCTC) will request information on your child’s hospital appointments (including inpatient, outpatient and A&E attendances) either directly from your NHS hospital and/or via the central NHS Digital system (NHS Digital records attendances at hospital in England only). Bangor University will not receive any identifiable information about you/your child.

Your child’s personal details, which will include their name, gender, NHS number, date of birth and postcode, will be shared with NHS Digital by LCTC in order for NHS Digital to provide information already in their possession on your child’s hospital attendances from 6 months before their entry into the study to when they leave the study. NHS Digital will then share this health information, which is regarded as a special category of information, with relevant members of the CASTLE Sleep-E research team at Bangor University. The data NHS Digital provides the CASTLE Sleep-E research team will have your child’s name, date of birth and other identifiers removed. At the end of the period specified in the NHS Digital Data Sharing Agreement, the data provided by NHS Digital will be securely destroyed.

For more information about how routine NHS data is collected and used please see the Appendix at the end of this document.

### 2.3 What are my choices about how my information is used?

You and your child can choose for your child to stop being part of the study at any time, without giving a reason, but we will keep information about you and your child that we already have. You can also choose to participate in the study but for us not to collect your NHS Digital information for the health economics analysis.

We need to manage your and your child’s research records, and the consent form you sign in specific ways for the research to be reliable. This means that we won’t be able to let you see or change the data we hold about you or your child.

### 2.4 Information sharing for other research

When you agree for you and your child to take part in a research study, the information about their health and care may be beneficial to researchers running other research studies in this organisation and in other organisations. These organisations may be universities, NHS organisations or companies involved in health and care research in this country or abroad. You and your child’s information will only be used by organisations and researchers to conduct research in accordance with the UK Policy Framework for Health and Social Care Research, or equivalent standards. You and your child’s information will be non-identifiable.

If you and your child agree for your child to take part in this study, you will have the option for you both to take part in future research using the data saved from this study.

### 2.5 Where can I find out more about how my and my child’s information is used?

You can find out more about how we use yours and your child’s information:

- at the study website: www.castlesleepetrial.org.uk
- at www.hra.nhs.uk/information-about-patients
- in the Health Research Authority leaflet available from www.hra.nhs.uk/patientdataandresearch
- by asking one of the research team
- by contacting the Data Protection Officers listed below;

King’s College London
King’s College Hospital
Bangor University (for matters relating to the NHS Digital subset –)
- by sending an email to or;
- by ringing us on 0151 794 8774

### 2.6 What if there is a problem?

If you have a concern about any aspect of this study, you should ask to speak to your study doctor who will do their best to answer your questions (details found one the first page of this document). If you remain unhappy and wish to complain formally, you can do this through the NHS Complaints procedure by contacting your local Patient Advice Liaison Service (PALS) office. Details of your local office can be obtained by asking your study doctor, GP, telephoning your local hospital or looking on the NHS choices website: http://www.nhs.uk/pages/home.aspx

Every care will be taken in the course of this study. However, in the unlikely event that you are injured by taking part, compensation may be available.

In the event that something does go wrong and you are harmed during the research and this is due to someone’s negligence then you may have grounds for a legal action for compensation against King’s College London and King’s College Hospital but you may have to pay your legal costs.

Regardless of this, if you wish to complain, or have any concerns about any aspect of the way you have been approached or treated by members of staff or about any side effects (adverse events) you may have experienced due to your participation in the study the normal National Health Service complaints mechanisms are available to you. Please ask your study doctor if you would like more information on this.

If you are concerned about how your information has been used during the trial, you can discuss this with the Information Commissioners Office (https://ico.org.uk/)

**Thank you for taking the time to read and consider this information sheet. Should you decide that you and your child can take part in the study, you will be given a copy of the completed consent form to keep.**

## Appendix

How routine (NHS Digital) hospital data is collected and used

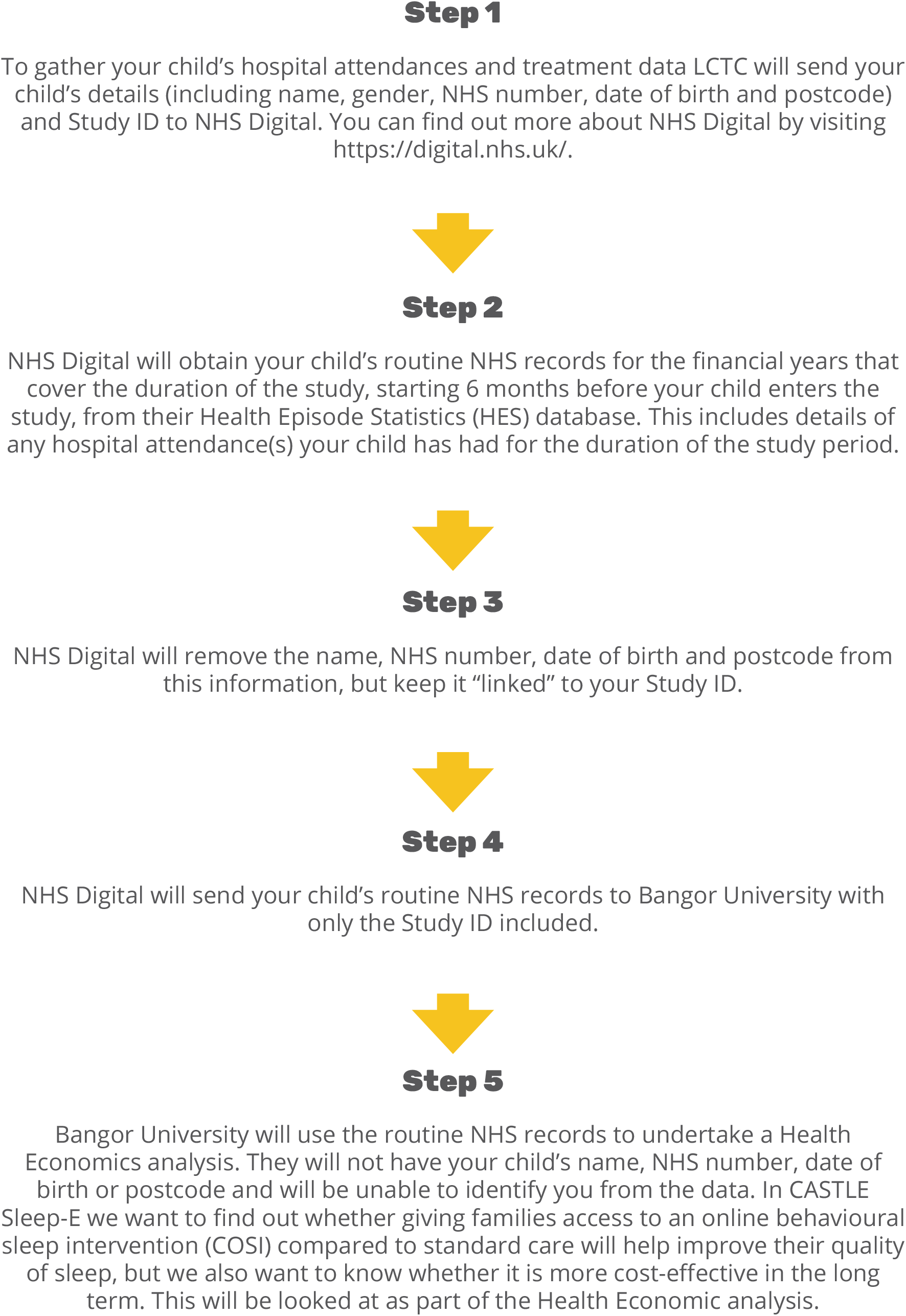

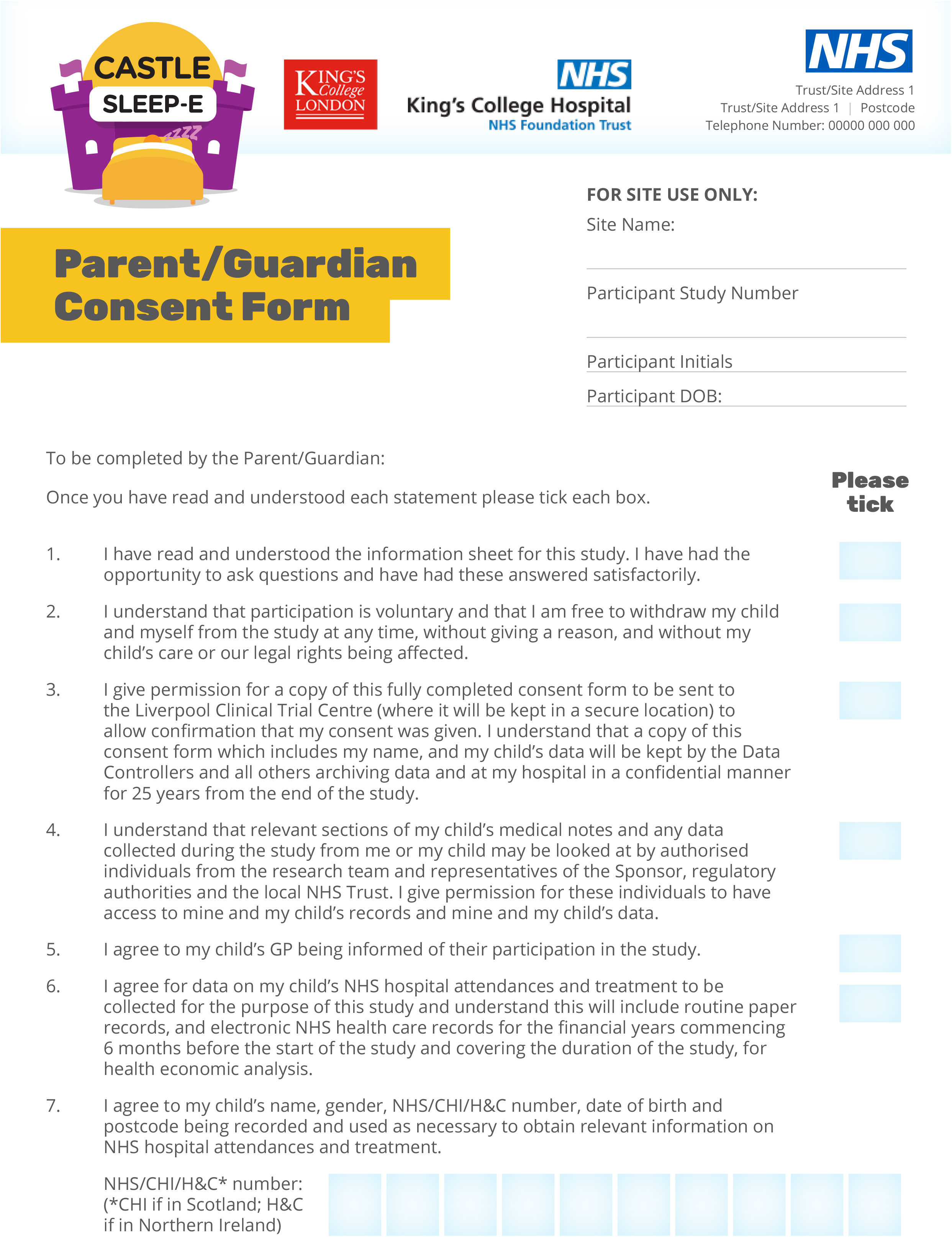

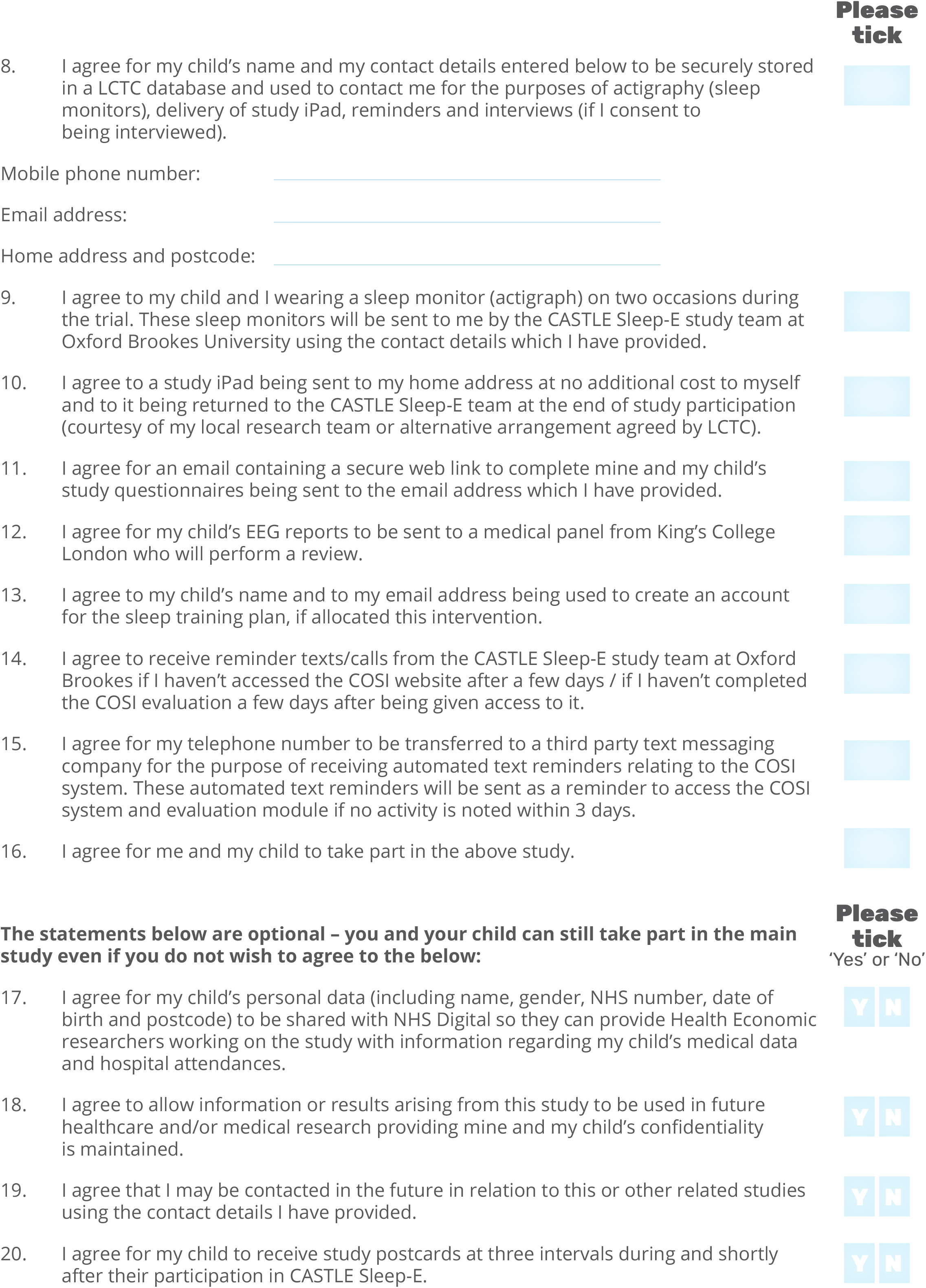

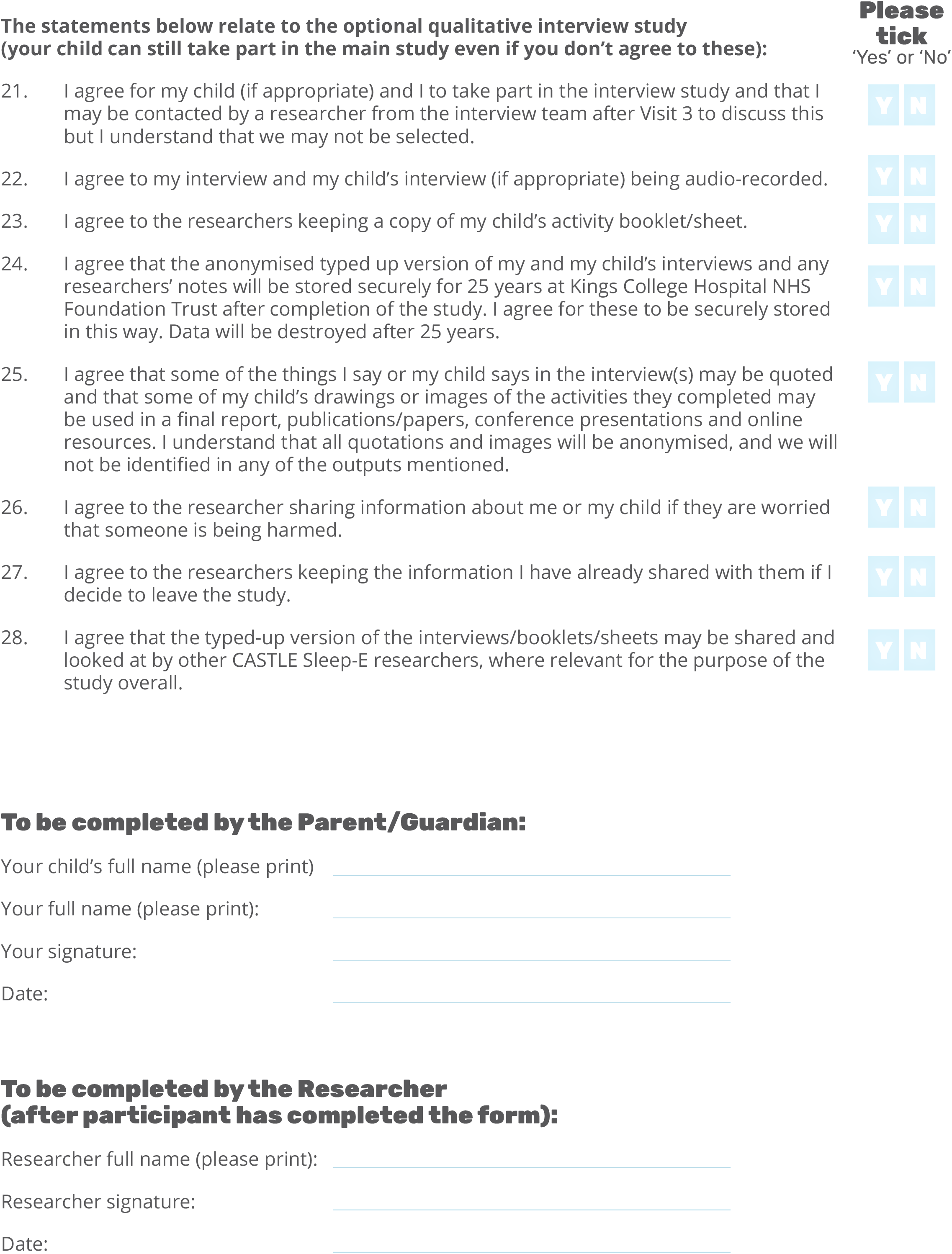

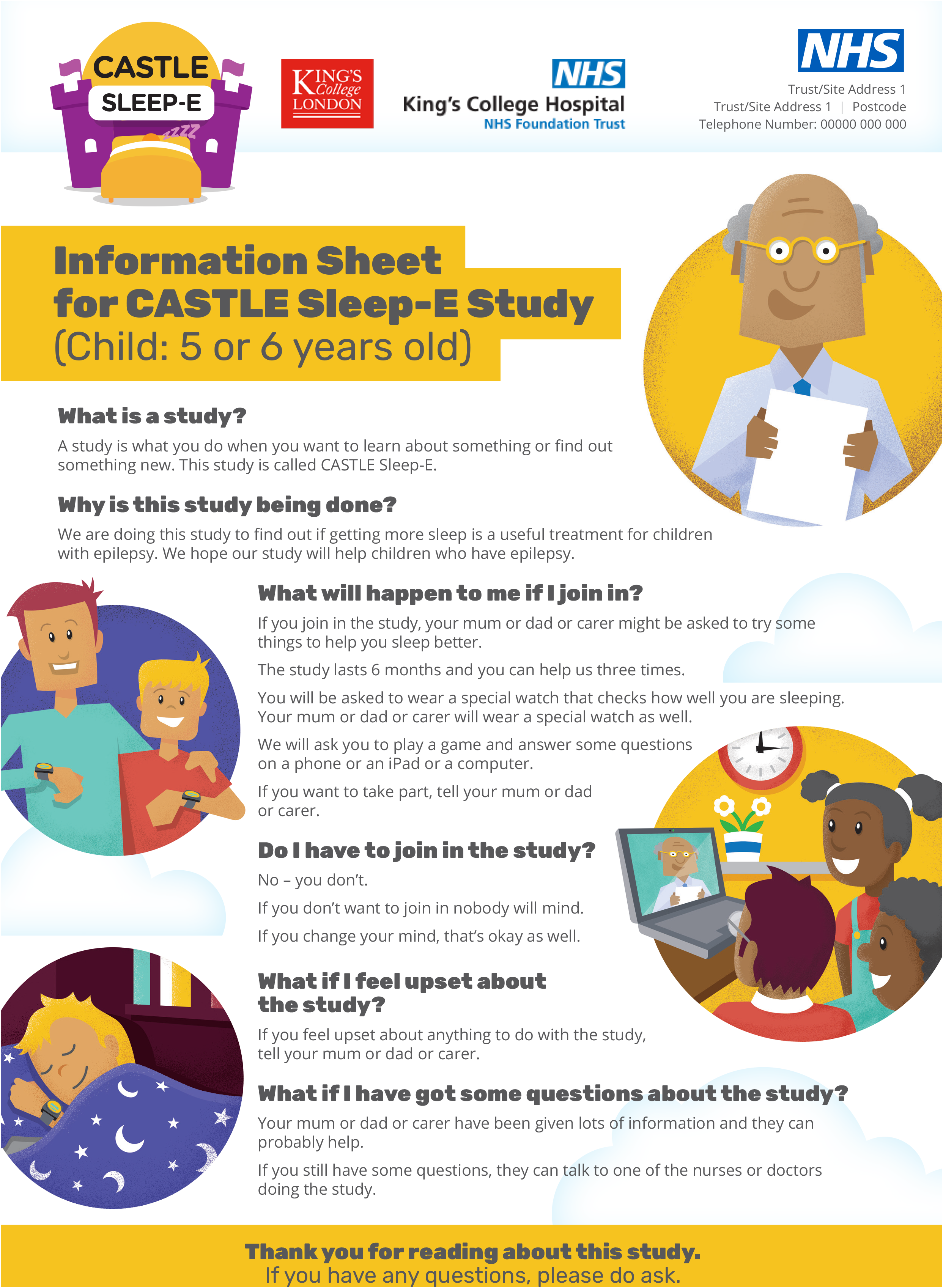

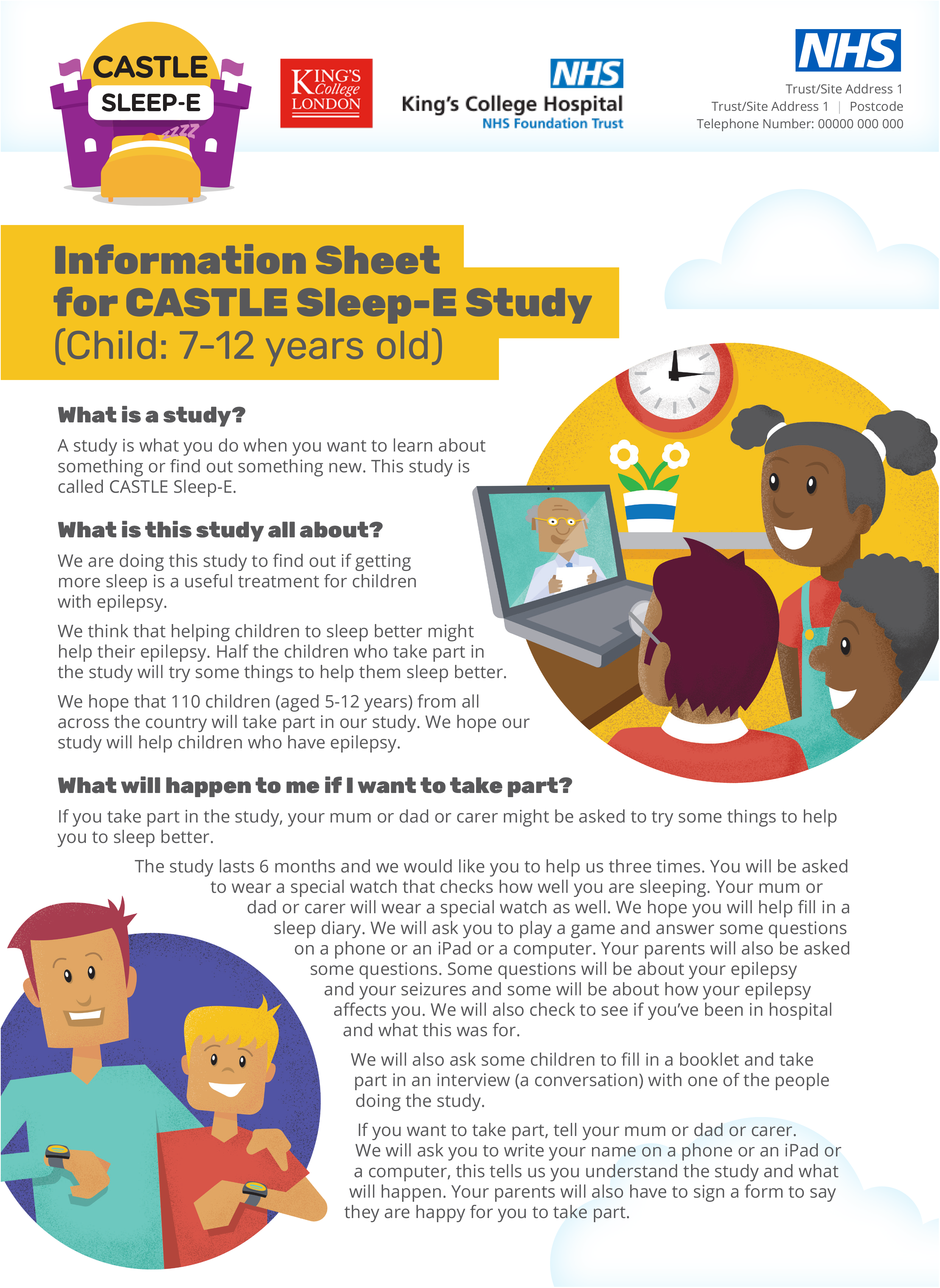

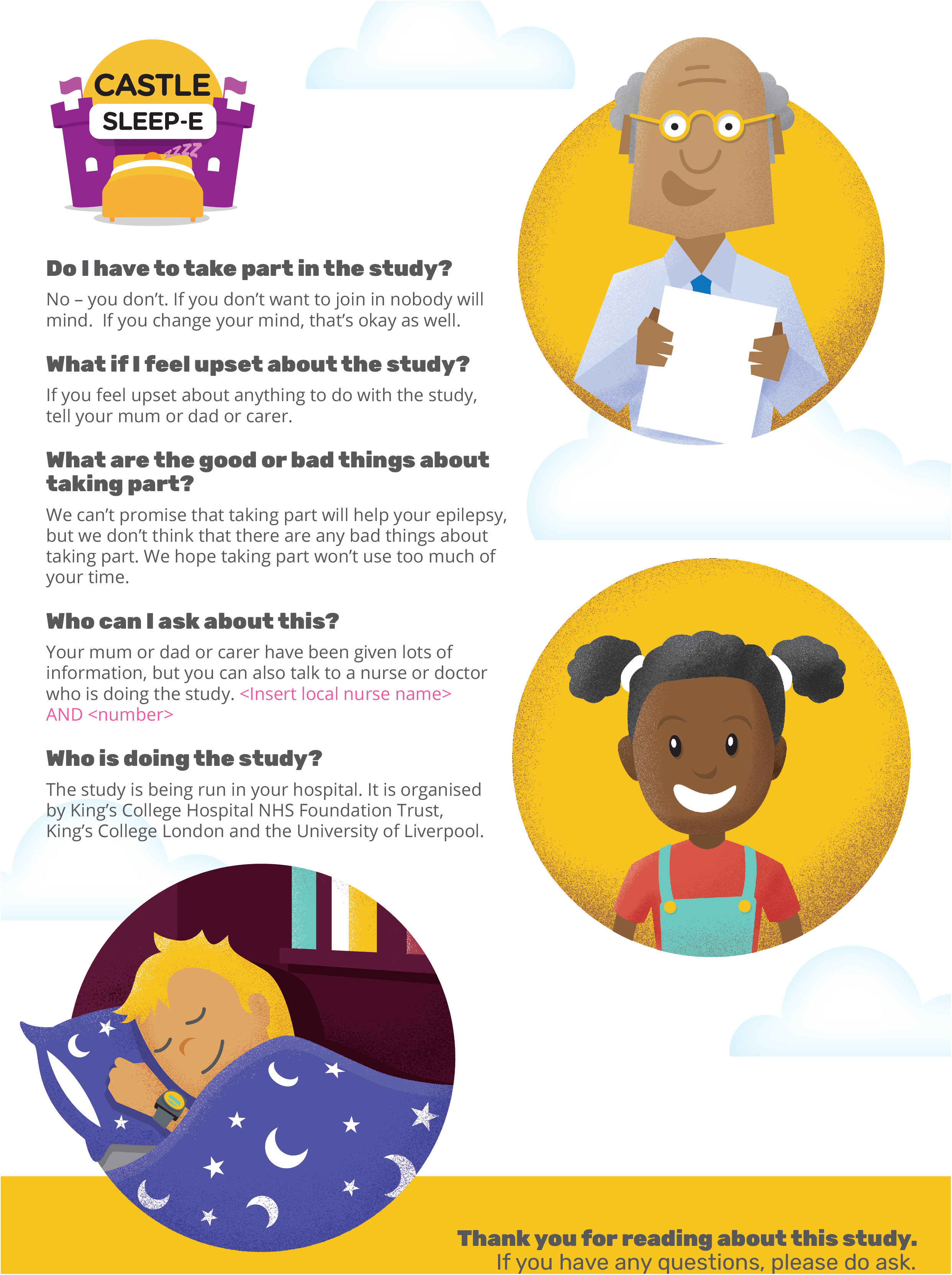

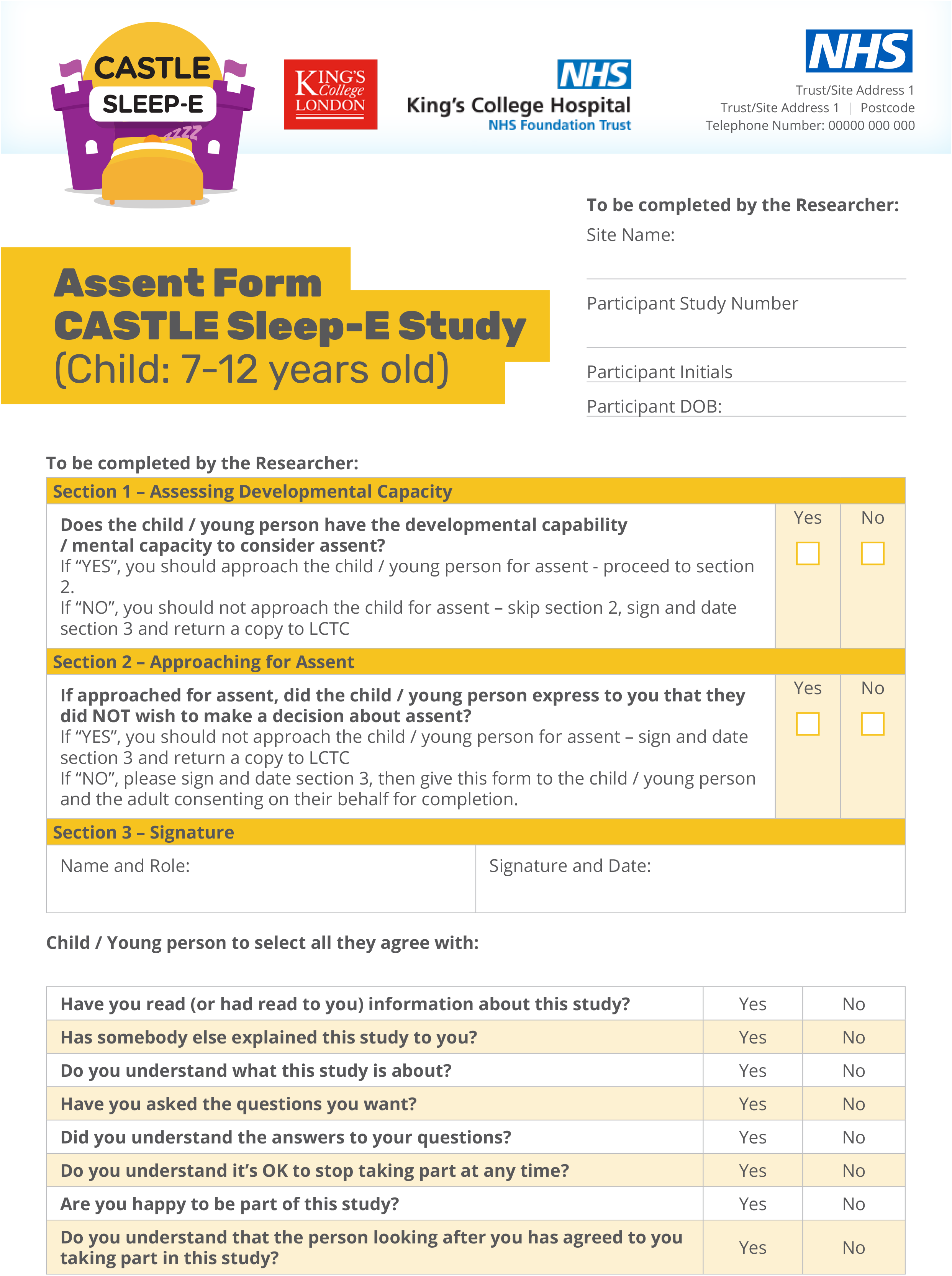

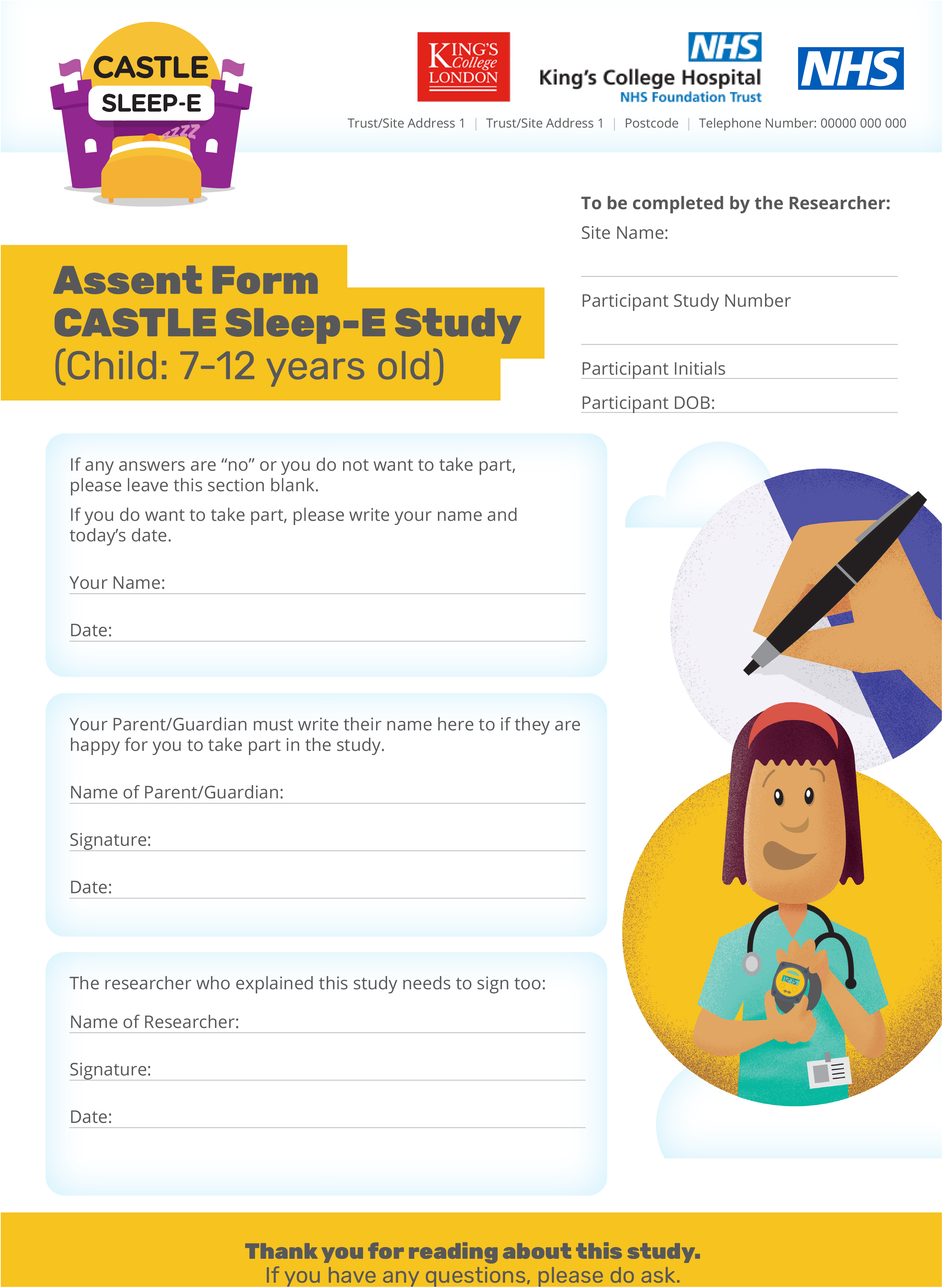

| Section/item | Item No | Description | Addressed on page number |
| --- | --- | --- | --- |
| **Administrative information** |  |  |  |
| Title | 1 | Descriptive title identifying the study design, population, interventions, and, if applicable, trial acronym | _____1_____ |
| Trial registration | 2a | Trial identifier and registry name. If not yet registered, name of intended registry | _____2_____ |
|  | 2b | All items from the World Health Organization Trial Registration Data Set | _Suppl. Table 1_ |
| Protocol version | 3 | Date and version identifier | _Suppl. Table 1_ |
| Funding | 4 | Sources and types of financial, material, and other support | 23, Suppl. Table 1 |
| Roles and responsibilities | 5a | Names, affiliations, and roles of protocol contributors | _____1, 23_____ |
|  | 5b | Name and contact information for the trial sponsor | _Suppl. Table 1_ |
|  | 5c | Role of study sponsor and funders, if any, in study design; collection, management, analysis, and interpretation of data; writing of the report; and the decision to submit the report for publication, including whether they will have ultimate authority over any of these activities | _____23_____ |
|  | 5d | Composition, roles, and responsibilities of the coordinating centre, steering committee, endpoint adjudication committee, data management team, and other individuals or groups overseeing the trial, if applicable (see Item 21a for data monitoring committee) | _Suppl. Table 3_ |

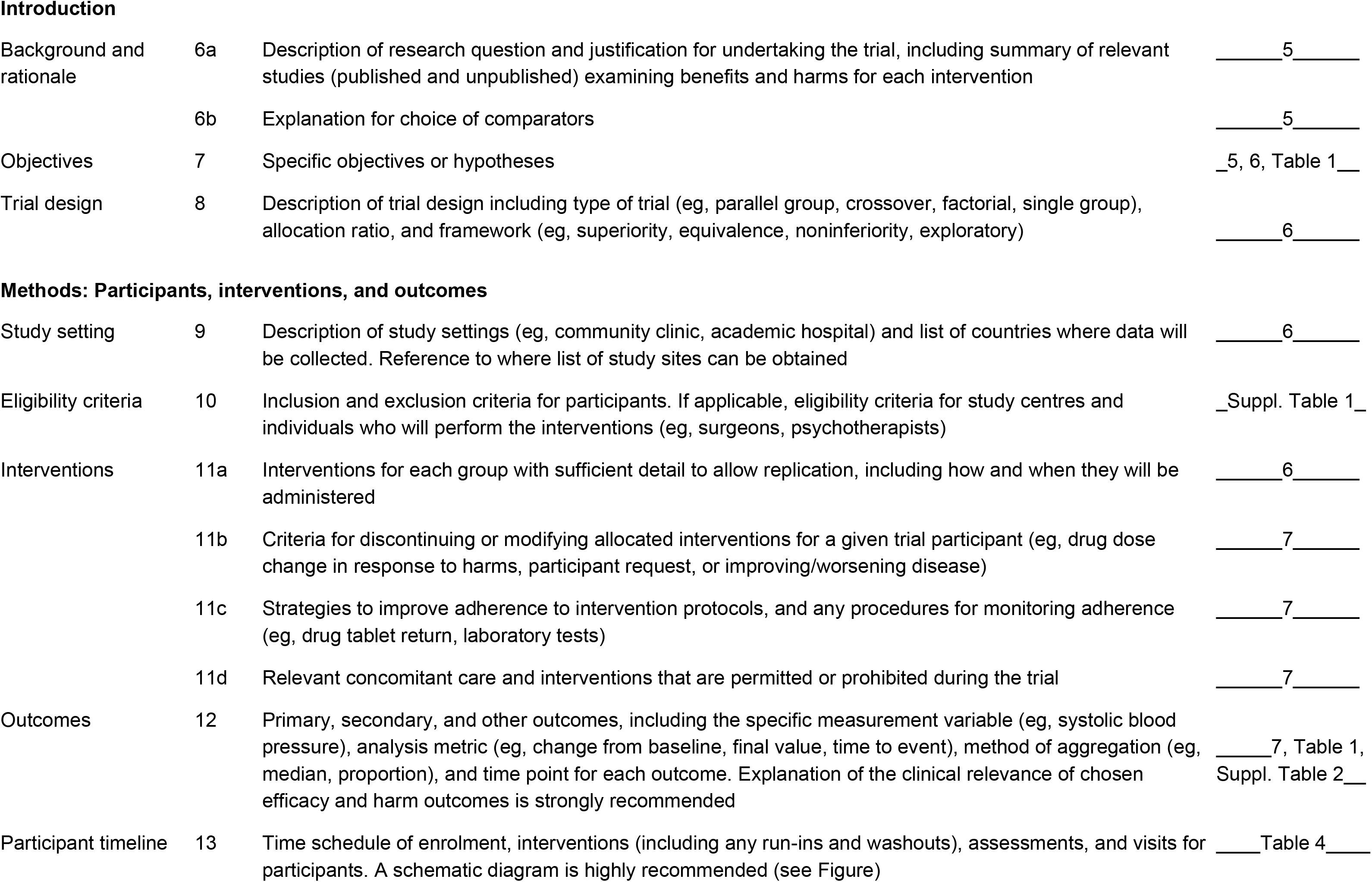

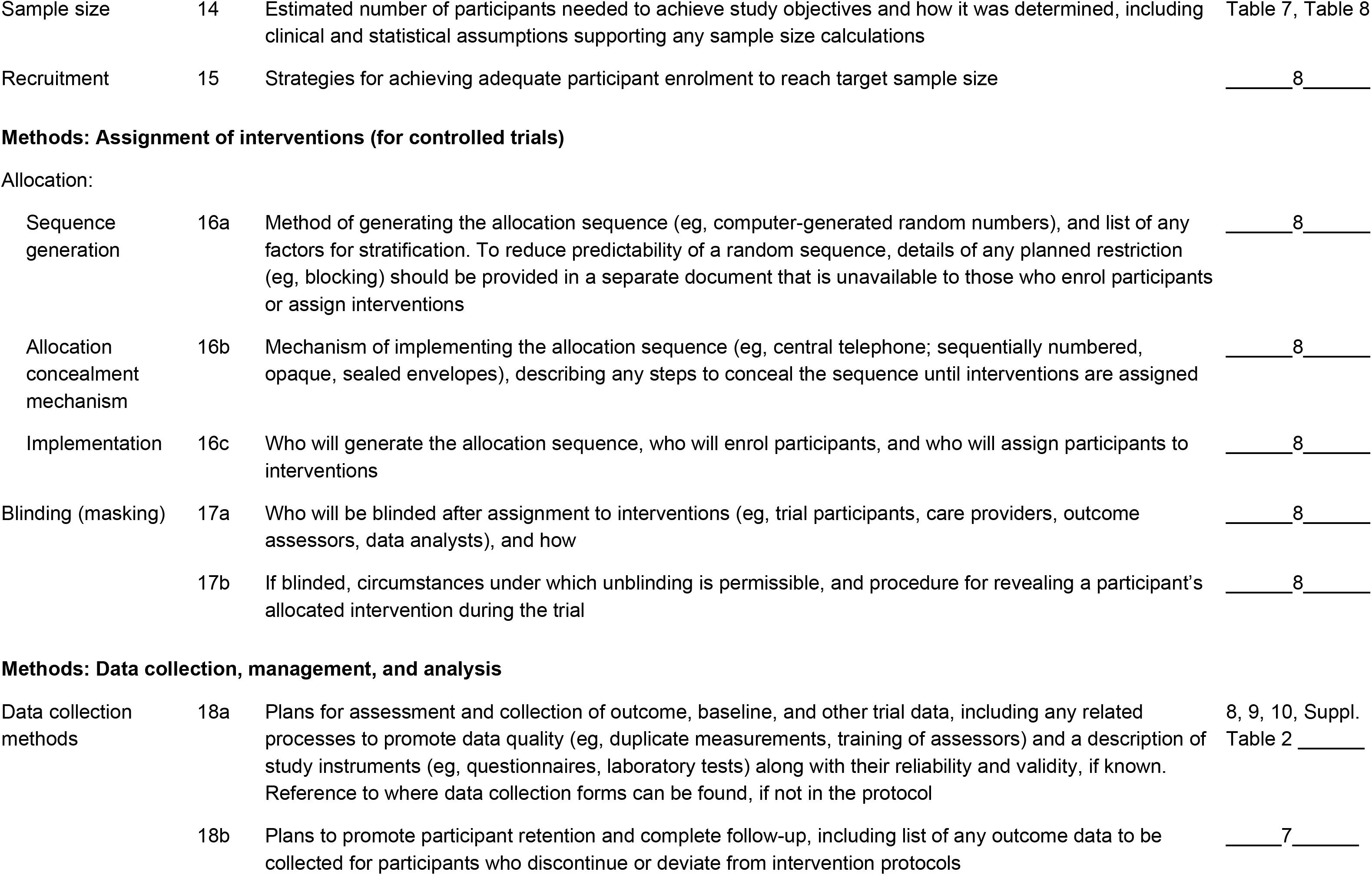

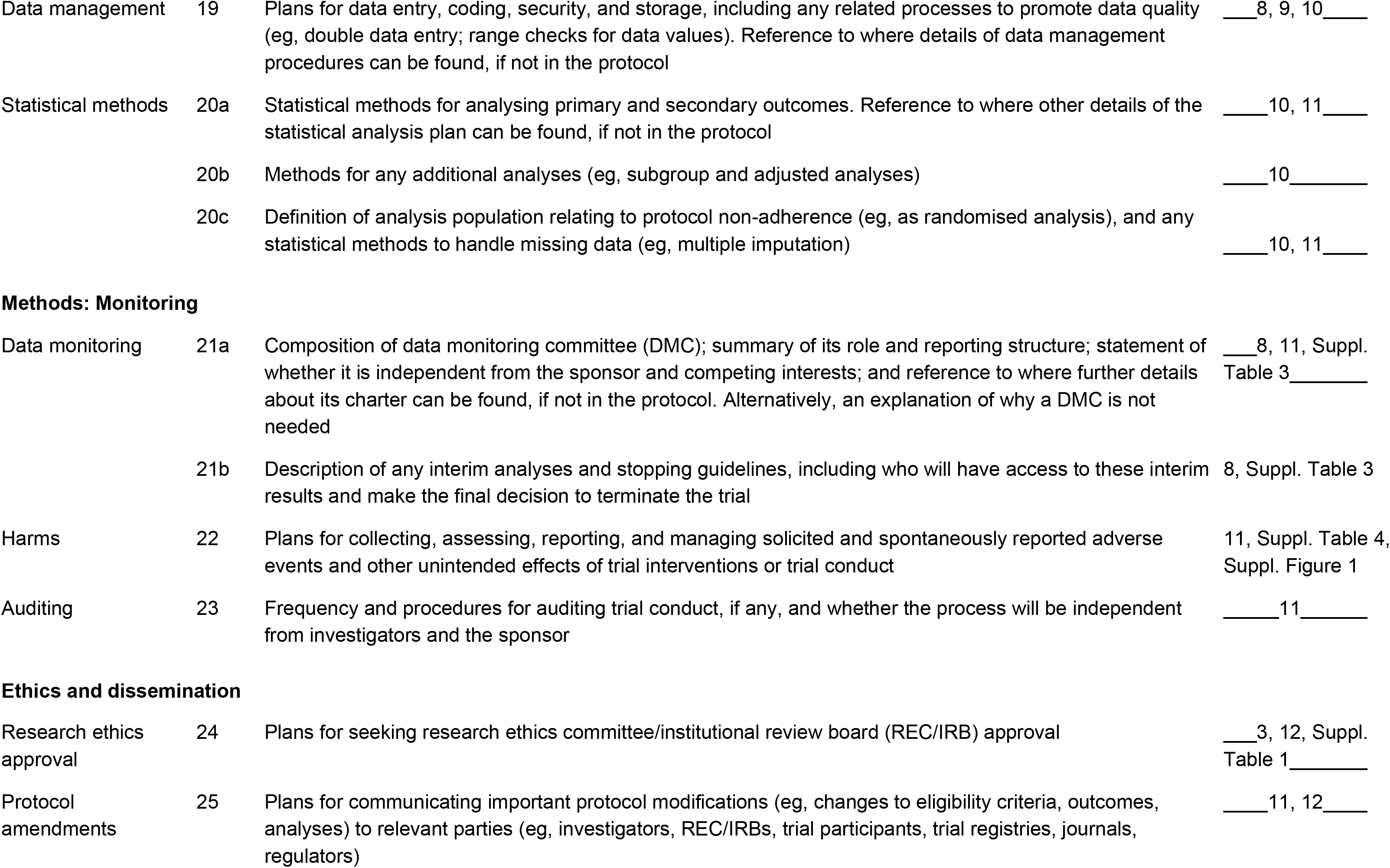

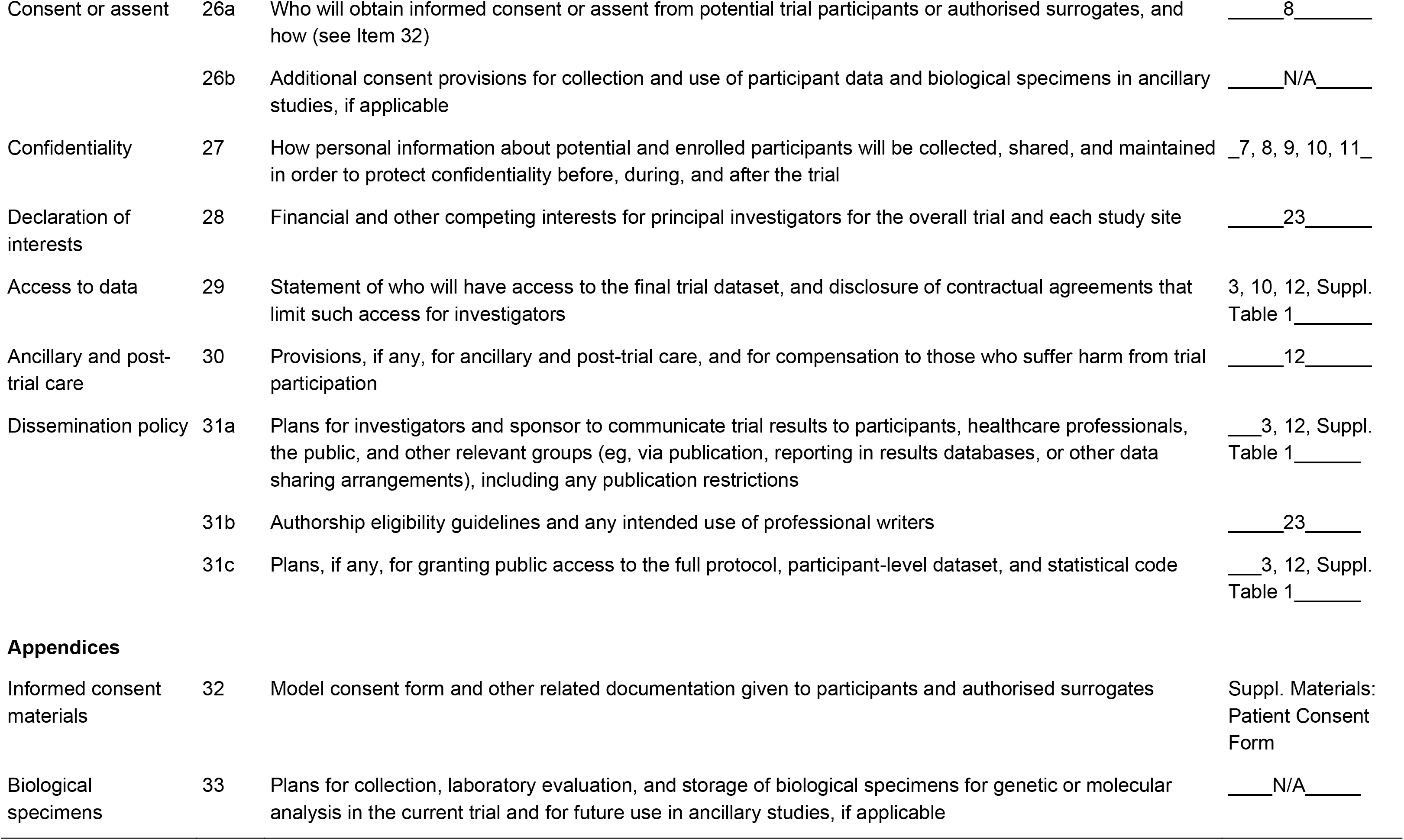

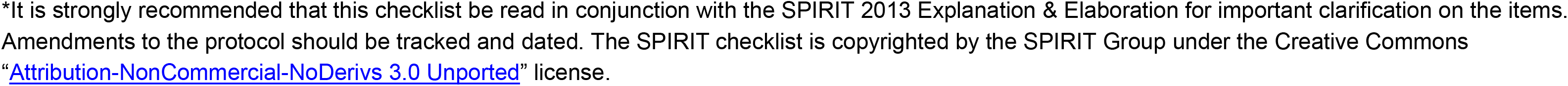

The SPIRIT-PRO Protocol Guidance Checklist
| Protocol Section | SPIRIT-PRO Item | Recommended Content | Page Addressed |
| --- | --- | --- | --- |
| **Administrative Information** |  |  |  |
| Roles and responsibilities | SPIRIT-5a-PRO Elaboration | Specify the individual(s) responsible for the PRO content of the trial protocol. | 23 |
| **Introduction** |  |  |  |
| Background and rationale | SPIRIT-6a-PRO Extension | Describe the PRO-specific research question and rationale for PRO assessment and summarize PRO findings in relevant studies. | Page 5, 7 |
| Objectives | SPIRIT-7-PRO Extension | State specific PRO objectives or hypotheses (including relevant PRO concepts/domains). | Page 7, Table 1, Table 5 |
| **Methods: Participants, Interventions, and Outcomes** |  |  |  |
| Eligibility criteria | SPIRIT-10-PRO Extension | Specify any PRO-specific eligibility criteria (eg, language/reading requirements or pre-randomization completion of PRO). If PROs will not be collected from the entire study sample, provide a rationale and describe the method for obtaining the PRO subsample. | Suppl. Table 1 |
| Outcomes | SPIRIT-12-PRO Extension | Specify the PRO concepts/domains used to evaluate the intervention (eg, overall health-related quality of life, specific domain, specific symptom) and, for each one, the analysis metric (eg, change from baseline, final value, time to event) and the principal time point or period of interest. | Table 1 |
| Participant timeline | SPIRIT-13-PRO Extension | Include a schedule of PRO assessments, providing a rationale for the time points, and justifying if the initial assessment is not pre-randomization. Specify time windows, whether PRO collection is prior to clinical assessments, and, if using multiple questionnaires, whether order of administration will be standardized. | Table 4 |
| Sample size | SPIRIT-14-PRO Extension | When a PRO is the primary end point, state the required sample size (and how it was determined) and recruitment target (accounting for expected loss to follow-up). If sample size is not established based on the PRO end point, then discuss the power of the principal PRO analyses. | Page 7-8 |

Calvert M, Kyte D, Mercieca-Bebber R, et al. Guidelines for Inclusion of Patient-Reported Outcomes in Clinical Trial Protocols: The SPIRIT-PRO Extension. *JAMA*. 018;319(5):483-494. doi:10.1001/jama.2017.21903
Note: The SPIRIT-PRO Extension should be used with the SPIRIT 2013 Statement and any other relevant SPIRIT Extensions, found at spirit-statement.org

The SPIRIT-PRO Protocol Guidance Checklist
| Protocol Section | SPIRIT-PRO Item | Recommended Content | Page Addressed |
| --- | --- | --- | --- |
| **Methods: Data Collection, Management, and Analysis** |  |  |  |
| Data collection methods | SPIRIT-18a(i)-PRO Extension | Justify the PRO instrument to be used and describe domains, number of items, recall period, and instrument scaling and scoring (eg, range and direction of scores indicating a good or poor outcome). Evidence of PRO instrument measurement properties, interpretation guidelines, and patient acceptability and burden should be provided or cited if available, ideally in the population of interest. State whether the measure will be used in accordance with any user manual and specify and justify deviations if planned. | Suppl. Table 2 |
|  | SPIRIT-18a(ii)-PRO Extension | Include a data collection plan outlining permitted mode(s) of administration (eg, paper, telephone, electronic, other) and setting (eg, clinic, home, other). | Page 8-10 |
|  | SPIRIT-18a(iii)-PRO Extension | Specify whether more than 1 language version will be used and state whether translated versions have been developed using currently recommended methods. | Suppl. Table 1 |
|  | SPIRIT-18a(iv)-PRO Extension | When the trial context requires someone other than a trial participant to answer on his or her behalf (a proxy-reported outcome), state and justify the use of a proxy respondent. Provide or cite evidence of the validity of proxy assessment if available. | Table 1, Table 4 |
|  | SPIRIT-18b(i)-PRO Extension | Specify PRO data collection and management strategies for minimizing avoidable missing data. | Page 7, 10 |
|  | SPIRIT-18b(ii)-PRO Elaboration | Describe the process of PRO assessment for participants who discontinue or deviate from the assigned intervention protocol. | Page 7 |
| Statistical methods | SPIRIT-20a-PRO Elaboration | State PRO analysis methods, including any plans for addressing multiplicity/ type I ($\alpha$) error. | Page 10 |
|  | SPIRIT-20c-PRO Elaboration | State how missing data will be described and outline the methods for handling missing items or entire assessments (eg, approach to imputation and sensitivity analyses). | Page 10-11 |
| **Methods: Monitoring** |  |  |  |
| Harms | SPIRIT-22-PRO Extension | State whether or not PRO data will be monitored during the study to inform the clinical care of individual trial participants and, if so, how this will be managed in a standardized way. Describe how this process will be explained to participants; eg, in the participant information sheet and consent form. | Page 8, 11 |

Calvert M, Kyte D, Mercieca-Bebber R, et al. Guidelines for Inclusion of Patient-Reported Outcomes in Clinical Trial Protocols: The SPIRIT-PRO Extension. *JAMA*. 018;319(5):483-494. doi:10.1001/jama.2017.21903
Note: The SPIRIT-PRO Extension should be used with the SPIRIT 2013 Statement and any other relevant SPIRIT Extensions, found at spirit-statement.org

